# Prenatal antidepressant exposure and longitudinal differences in body mass index up to 8 years of age in the offspring born to mothers with pre-pregnancy depressive and/or anxiety in the Norwegian Mother, Father and Child Cohort Study

**DOI:** 10.1101/2024.07.09.24310139

**Authors:** Nhung TH Trinh, Sina Rostami, Michele Pedroncelli, Rosa Cheesman, Per Magnus, Stefan Johansson, Ole A. Andreassen, Angela Lupattelli

**Author notes:** **Corresponding author**: Nhung TH Trinh; Post box 1068 Blindern, 0316 Oslo, Norway;.

## Abstract

**Importance:** No study with available data from birth into late childhood has explored how prenatal antidepressant exposure affects offspring body mass index (BMI) throughout childhood.

**Objective:** To determine the association between prenatal antidepressant exposure and longitudinal differences in child BMI up to age 8 years.

**Design, Setting, and Participants:** We used data from the Norwegian Mother, Father, and Child Cohort Study (MoBa) linked to the Medical Birth Registry of Norway and the MoBa Genetics. We included 6,084 pregnancy-child dyads (singleton, liveborn) with available parent-reported data on child BMI from birth up to 8 years of age, born to women with depression/anxiety prior to pregnancy. Analysis was performed between January 2023 and April 2024.

**Exposures:** Prenatal antidepressant exposure was categorized as i) *continued* antidepressants in pregnancy (n=626); ii) *discontinued* antidepressants proximal to pregnancy (n=412); or iii) *unexposed* to antidepressants both before and during pregnancy (n=5,046).

**Main outcomes and measures:** Child BMI up to 8 years of age. Mean BMI differences over time across antidepressant exposure groups were compared using multilevel mixed-effect linear models.

**Results:** Children born to mothers who continued antidepressant into pregnancy had comparable childhood BMIs with those born to unexposed mothers or mothers who discontinued antidepressant proximal to pregnancy. Higher BMI was observed up to 3 years of age among male offspring born to antidepressant continuers compared to discontinuers, especially in those exposed to selective-serotonin-reuptake-inhibitor before pregnancy (mean difference in BMI, β=0.334; 95% CI: 0.081 to 0.588 at baseline). Lower BMI was seen among female offspring born to continued vs. discontinued mothers and the gap became larger over time, especially between low-moderate use of antidepressant vs. discontinuation during pregnancy. Analyses integrating parental genetic liability for depression, BMI, and antidepressant response using polygenic risk scores in a sub-population (n=1,913) suggests potential influence of the genetic component on the differences in BMI across antidepressant trajectory groups in some strata.

**Conclusion and relevance:** The longitudinal childhood BMI of children born to mothers with pre-pregnancy depression/anxiety did not differ across prenatal antidepressant exposure trajectories. Exploratory analyses revealed differences at specific time frames which might be sex-specific and potentially influenced by genetic liability profiles.

**KEY POINTS:** *Question:* Does prenatal antidepressant exposure affect longitudinal childhood BMI?

*Findings:* In this cohort study of 6084 pregnancy-child dyads in mothers with pre-pregnancy depressive/anxiety disorders, no difference in longitudinal childhood BMI across prenatal antidepressant exposure groups were observed. Exploratory analyses revealed differences at specific time frames which might be sex-specific and potentially influenced by parental genetic liability profiles.

*Meaning:* Longitudinal BMI throughout childhood of children born to mothers with pre-pregnancy depression/anxiety did not differ across prenatal antidepressant exposure trajectories. Further research is needed to investigate the time-dependent, sex-specific, and genetic-related aspects of some strata of antidepressant exposure on BMI differences.

## INTRODUCTION

Antidepressants constitute the mainstay pharmacotherapy of depression, anxiety, and other related disorders in both pregnant women and the general population.^1^ It is estimated that 7-15% of pregnant women with depressive/anxiety disorders receive antidepressant during pregnancy.^2^ A sub-optimally treated psychiatric illness during pregnancy may result in poor pregnancy/obstetric outcomes, inadequate nutrition and increased substance use.^3^ However, uncertainties about the safety of *in utero* exposure to antidepressants for the offspring lead to high rate of antidepressant discontinuation among the pregnant population.^4,5^ Establishing the safety profile of prenatal antidepressant exposure including both immediate and long-term impact on the offspring, is crucial to support the evidence-based decision making in the management of psychiatric illnesses in pregnant women.

Both underlying psychiatric illnesses and antidepressant use during pregnancy could alter the development of the serotonergic system in early fetal life.^6,7^ Serotonergic alterations might lead to several long-term metabolic consequences in the offspring (e.g., body weight and other related health correlates and school performance).^8,9^ However, little is known about the role of prenatal antidepressant exposure in these associations. Some studies suggested that prenatal antidepressant exposure has the potential to affect growth and body weight in the offspring with sex-specific results.^10–12^ Specifically, Grzeskowiak et al. found a 78% greater risk for childhood overweight with prenatal antidepressant exposure among males, but not among females. In addition, prenatal antidepressant exposure is associated with increased risk of low birth weight in the infant and prematurity, which can in turn affect growth and metabolic health later in life.^13^ The current evidence on this research question is limited with mixed results, only focused on childhood overweight at specific time points (i.e., 4-5 years and 7 years of age), did not take duration of prenatal antidepressant exposure nor the impact of genetic predispositions and familial environment into consideration.^14^

This present study sought to examine the influence of prenatal antidepressant exposure on BMI throughout childhood in children born to mothers with pre-pregnancy depressive/anxiety disorders considering treatment duration and parental genetic liability for depression and BMI.

## METHOD

### Data sources

This study was based on data from the Norwegian Mother, Father, and Child Cohort Study (MoBa), linked with MoBa Genetics and the Medical Birth Registry of Norway (MBRN) using unique project-specific individual IDs generated from the personal identification number assigned at birth or immigration to Norway.^15^

MoBa is a prospective population-based pregnancy cohort study conducted by the Norwegian Institute of Public Health. The cohort comprises more than 100,000 pregnant women from all over Norway, recruited during the period between 1999 and 2008 with participation rate of about 41%. In MoBa, all women who consented to participate received three questionnaires during pregnancy at gestational weeks 17 (Q1), 22 (Q2), and 30 (Q3) and multiple postnatal questionnaires when the child was 6 months (Q4), 18 months (Q5), three years (Q6), five years (Q7), seven years (Q8), eight years (Q9) and so on. Questionnaires Q1 and Q3 collected a wide range of information on socio-demographic characteristics, outcomes of previous pregnancies, medical history, maternal health, lifestyle habits, and medication exposures as well as other exposures during pregnancy.^15^

Blood samples were collected from both parents during pregnancy (proximal to gestational week 18) and from the mothers and children (umbilical cord) at birth. Genetic data was retrieved from MoBa Genetics v1.0 which was based on a multi-year effort across various research projects, employing diverse selection criteria, genotyping platforms, and genotyping centres.^16,17^ The current MoBa genetics dataset comprises imputed genetic data for 98,110 individuals (∼32,000 parent offspring trios), derived from nine batches of participants, who make up four study cohorts. Information on MoBa Genetics study cohorts, quality control and imputation is available online https://github.com/folkehelseinstituttet/mobagen.^18^ There were post-imputation quality control procedures, and eventually 93,582 individuals and 6,797,215 SNPs were included. Further details on quality control procedures and post-imputation quality control for MoBa Genetics was previously described.^19^ The existence of genotyping data was not a prerequisite for inclusion in the study, rather we did subsequent analyses in subsample of participants we had genetics data on.

Polygenic risk score (PRS) was calculated based on summary statistics from recent genome-wide association studies in European populations for depression, BMI and antidepressant response.^20–22^ For depression and BMI, we fit PRS based on different combinations of SNPs at different GWAS p-value thresholds and used the most predictive combination of SNPs as PRS fitted for depression for 5354 mothers (thereafter called “maternal depression PRS”, 221,739 SNPs), BMI for 5526 mothers (thereafter called “maternal BMI PRS”, 29,414 SNPs), and BMI for 2237 fathers (thereafter called “paternal BMI PRS”, 50,318 SNPs). For anti-depressant response, given that there are currently not many published GWAS with large sample sizes, our PRS was based on 27 SNPs which had GWAS p-value <5.005e-05. More details are given in **Appendix 1**.

The MBRN was established in 1967 and is based on compulsory notification of all live births, stillbirths, and induced abortions after week 12 in Norway. The registry comprises maternal medical records during prenatal care, as well as mother and child health at the delivery ward.^23^

### Study population

We included singleton pregnancies that resulted in livebirth in women who self-reported depressive and/or anxiety disorders before pregnancy in the Q1 questionnaire, complemented with diagnostic records from MBRN (ICD-10 codes: F32, F33, F40, F41). In Q1, women were presented with a list of long-term conditions, including depressive and anxiety disorders. To obtain reported information on medication exposure throughout pregnancy, we also required that Q3 questionnaire was returned. We excluded pregnancies with (i) no information on BMI throughout the follow-up period (i.e., from 6 weeks of age to 8 years old) or (ii) unknown timing of antidepressant exposure, and (iii) antidepressant initiation during pregnancy due to small sample size of this group.

### Antidepressant exposure groups

Information about type and timing of antidepressant exposure in pregnancy was retrieved from the Q1 and Q3 questionnaires. Women were asked to report the type of medications and the duration of use in the six months before pregnancy as well as during pregnancy The timing of use could be reported by monthly intervals (e.g., from week 0-4, 5-8, through 13+ in Q1, and from week 13-16 through week 29+ in Q3). We categorized antidepressants into selective serotonin reuptake inhibitors (SSRIs, ATC code N06AB), serotonin and norepinephrine reuptake inhibitors (SNRIs, ATC codes N06AX16 and N06AX21), tricyclic antidepressants (TCAs, ATC code N06AA) and other antidepressants (OADs, i.e., ATC codes N06AX03, N06AX06, N06AX11, N06AX12 and N06AX18).

Based on the reported antidepressant exposure before and during pregnancy, we classified our population into: (1) *continued antidepressant in pregnancy* – pregnancy-child dyads in women who reported using antidepressants in the six months before and anytime during pregnancy, (2) *discontinued antidepressant before pregnancy* – pregnancy-child dyads in women who reported using antidepressants before but not during pregnancy, (3) *antidepressant unexposed* – pregnancy-child dyads in women who reported not using antidepressant neither before nor during pregnancy.

Among those who continued antidepressant treatment during pregnancy, we further classified them as: *antidepressant sustained use* (> 5 monthly intervals), *low-moderate use* (2-5 monthly intervals), and *antidepressant discontinuers in pregnancy* (1 monthly interval only). We explored the potential effect of antidepressant duration of exposure during pregnancy by comparing *sustained use* and *low-moderate use* vs. *antidepressant discontinuers in pregnancy*.

### Child anthropometric data

Postnatal anthropometric data of the children were reported in the MoBa questionnaires by the mothers at 11 time points and in six different questionnaires: around the age of 6 weeks, 3 months and 6 months (Q4), around the age of 8 months, 1 year and 18 months (Q5), around the age of 2 years and 3 years (Q6), the age of 5 years (Q7), the age of 7 years (Q8), and 8 years (Q9). From 6LJweeks to 18LJmonths, mothers were asked to refer to their child’s health card (measurements performed by nurses), whereas no specification was provided for measurements from 2 to 8LJyears. We computed BMI as weight in kilograms divided by height in meters squared and actual BMI values were used as outcome in all analyses.

We used the following criteria from the Bergen study, a nationwide study in Norway assessing growth charts in children in 2003-2006, to omit values that would be biologically implausible: 1) a z-score for weight less than −6 or greater than 5 using the Bergen study percentiles, 2) a z-score for length less than −6 and greater than 6 using the Bergen study percentiles.^24^

### Covariates and handling of missing data

Based on previous knowledge and a directed acyclic graph approach, we selected the following covariates for model adjustment: sociodemographic-lifestyle-reproductive factors before pregnancy (i.e., maternal age, parity, marital status, obstetric comorbidity index adapted from Bateman et al.,^25^ maternal education, maternal gross yearly income, smoking and alcohol use, BMI at conception, history of abortions/miscarriages, folic acid intake, paternal age, paternal education, paternal social benefits, and paternal BMI), maternal psychiatric correlates (i.e., Lifetime History of Major Depression measured in Q1), comedication before pregnancy (i.e., opioid analgesics – ATC code N02A, benzodiazepines/z-hypnotics - ATC codes N05B and N05C, antipsychotics and mood stabilizers - ATC code N05A, and antiepileptics - ATC code N03A). This set of covariates was used in all models. Confounding adjustment was done via stabilized inverse probability of treatment weighting (SIPTW) using propensity score, trimmed at 1^st^ and 99^th^ percentiles.^26^

We first investigated the patterns of missing data of confounding variables. Under the assumption that data were missing at random, multiple imputation (n=10) of missing values was performed via chained equations with 100 iterations. The imputation includes exposure and outcome variables, maternal age, parity, co-medication, pregnancy outcome, mother identity number, maternal illnesses, smoking, alcohol, and illicit substances abuse accounting for sex-differences in BMI (option *augment* by child sex in STATA).

### Statistical analyses

We first descriptively analysed the study population characteristics across antidepressant exposure groups. Second, we fitted three-level (occasions of child’s assessment, pregnancy-child dyad, mother) mixed-effects linear spline regression using full information maximum likelihood and an unstructured covariance with a random intercept (levels 2 and 3) and a random slope (level 2) to estimate differences in BMI across antidepressant exposure groups over time. We conducted the following comparisons: *continued antidepressant during pregnancy* vs *unexposed*, *continued antidepressant during pregnancy* vs *discontinued antidepressant before pregnancy*, *sustained use* vs. *discontinuers in pregnancy*, *low/moderate use* vs *discontinuers in pregnancy*. Time (i.e., child’s age in years) was scaled for postnatal questionnaire completion date and modelled as continuous. Linear splines with knot points were positioned at 4, 8, and 18 months. Adjusted models included an interaction term between time and antidepressant exposure, child’s age at baseline, and trimmed stabilized inverse probability of treatment weights as fixed effects. The crude and adjusted beta coefficients (β) with 95% confidence interval (CI) represent the mean difference in BMI outcomes between prenatal antidepressant exposure groups.

### Additional analyses

First, because some mothers may contribute more than one pregnancy in the final study population, we repeated the analyses in first pregnancies to explore how dependencies among observations affect the study findings. Second, we repeated the analyses in the term born children to explore the potential impact of prematurity on the outcome of interest. Third, we repeated the analyses in children with non-low birth weight to explore how weight at birth affects longitudinal differences in BMI later in life. Fourth, measured time-varying confounders (i.e., SCL-5 scores measured by 5/25 items in the Hopkins Symptoms Checklist in Q1 and Q3)^27^ were not included directly in the calculation of SIPTW. We included these covariates in the linear mixed models to further control for depressive symptoms severity during pregnancy. Fifth, we restricted our population to pregnancies exposed to SSRIs in continuers and discontinuers, as SSRIs are the most commonly used antidepressants for pregnant women and these drugs selectively inhibit the reuptake of the serotonin, which is the neurotransmitter involved in weight regulation.^10^ Sixth, we attempted to explore whether shared genetic influences could confound the relationship between prenatal antidepressant use and longitudinal BMI across childhood using a subsample for whom we had genetics data (1,913 pregnancy-child dyads).^28^ For this purpose, we adjusted for the polygenic risk score (PRS) fitted for maternal depression, maternal BMI, paternal BMI, and PRS based on 27 SNPs for antidepressant response in continuous form and z-scores as well as genotyping batch groups (n=6), and the five first principal components to account for population substructure parental genetic liability profiles might be helpful to identify strata with higher risks for abnormal childhood BMI. We then performed several stratified analyses by selecting participants in the upper or lower quartiles of PRSs for maternal depression, maternal BMI, paternal BMI, and antidepressant response. Details about calculation of PRSs were summarized in **Appendix 1**.^20,21^

Data management and analyses were performed using STATA 17 statistical software (Stata Corporation, College Station, TX).

### Ethical approval

The establishment of MoBa and initial data collection was based on a license from the Norwegian Data protection agency and approval from The Regional Committees for Medical and Health Research Ethics. The MoBa cohort is based on regulations in the Norwegian Health Registry Act. The current study was approved by The Regional Committees for Medical and Health Research Ethics on 12th February 2020 (reference number: 63566/REK Sør-Øst), and by the Norwegian Data protection agency (reference number: 672954).

## RESULTS

A total of 6,084 children were included in the study (**Figure 1**). Overall, 626 (10.3%) women were defined as *continued antidepressant* in pregnancy, 412 (6.8%) were *discontinued antidepressant before pregnancy* while 5,046 (82.9%) remained *unexposed*. Among *continued antidepressant mothers*, there were 173 (27.6%) with *sustained use*, 213 (34.0%) with *low-moderate use* and 240 (38.3%) *discontinued antidepressants during pregnancy*. SSRIs were the most used antidepressants (up to 80%) followed by SNRIs (about 10%) both before and during pregnancy, and mainly as monotherapy. General characteristics of the study population according to antidepressant exposure groups are presented in **Table 1 and Table S1**. *Mothers who continued antidepressant during pregnancy* were more likely to use alcohol and illicit substances before pregnancy, have co-medication before pregnancy, have history of major depression, and have more severe depressive/anxiety symptoms compared to the other groups of exposure. Differences in the distribution of characteristics across antidepressant exposure groups were well-balanced after weighting (see **Figure S1 and S2**).

**Figure 1.**
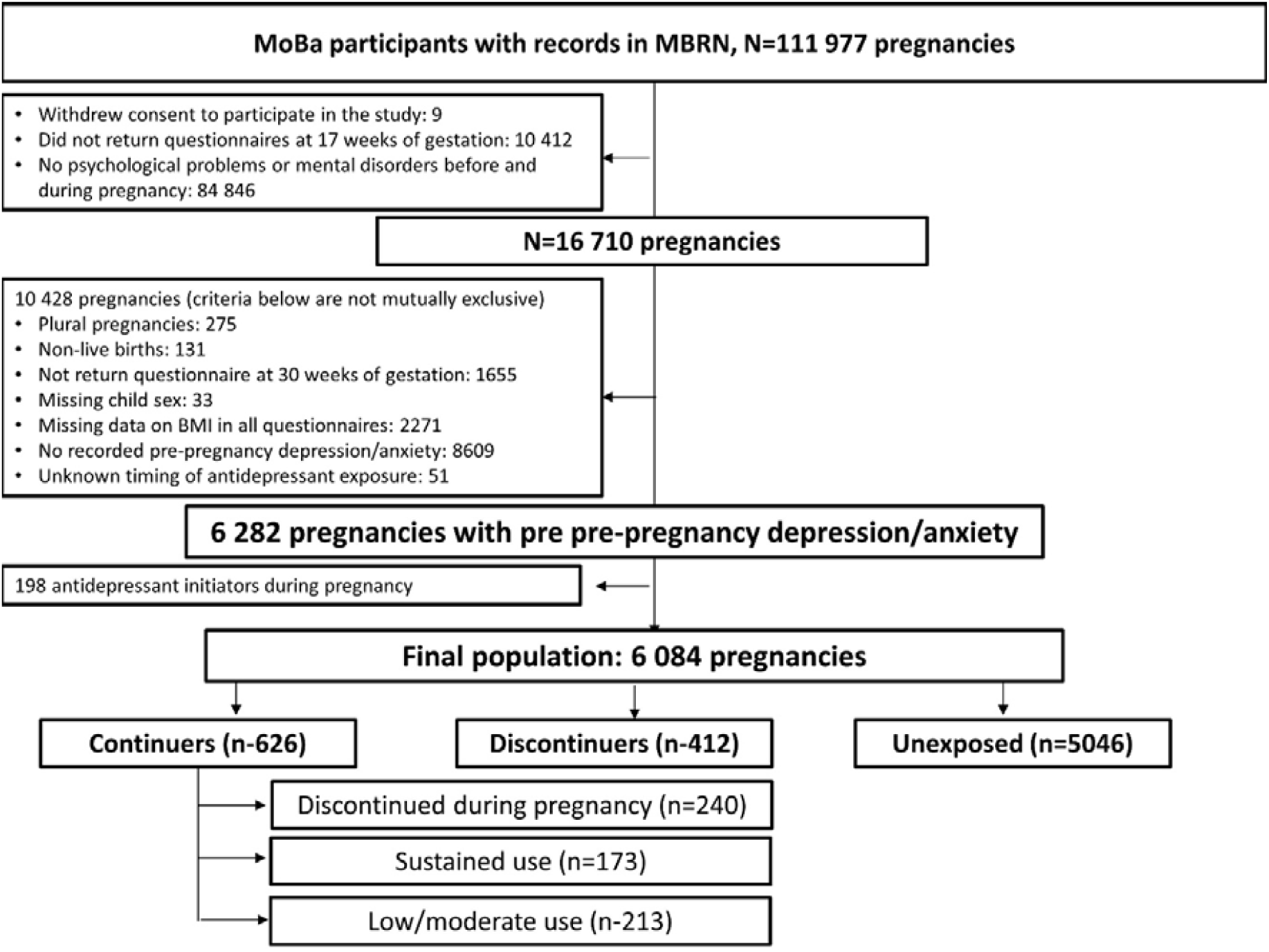
Flowchart of study population

**Table 1.**
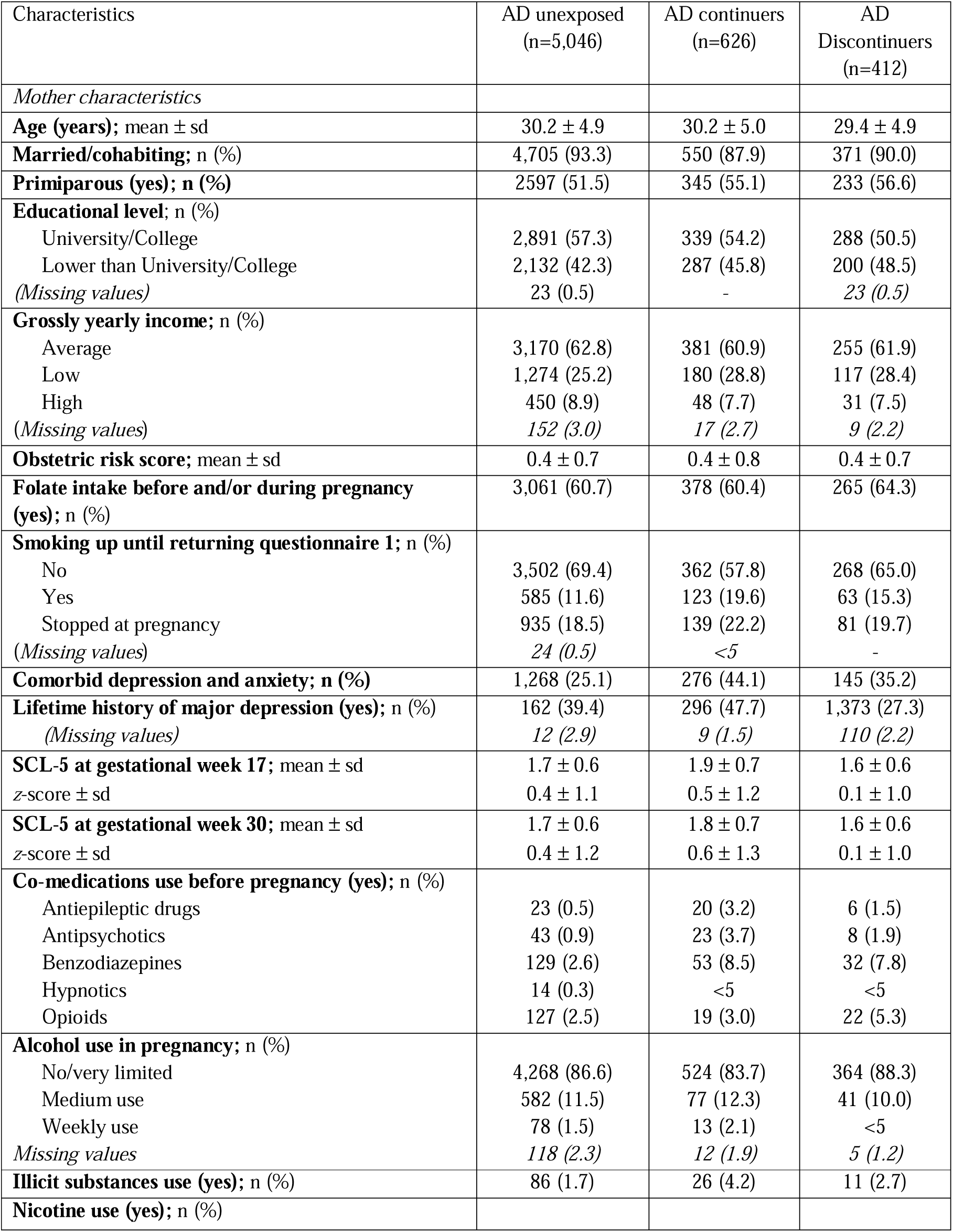
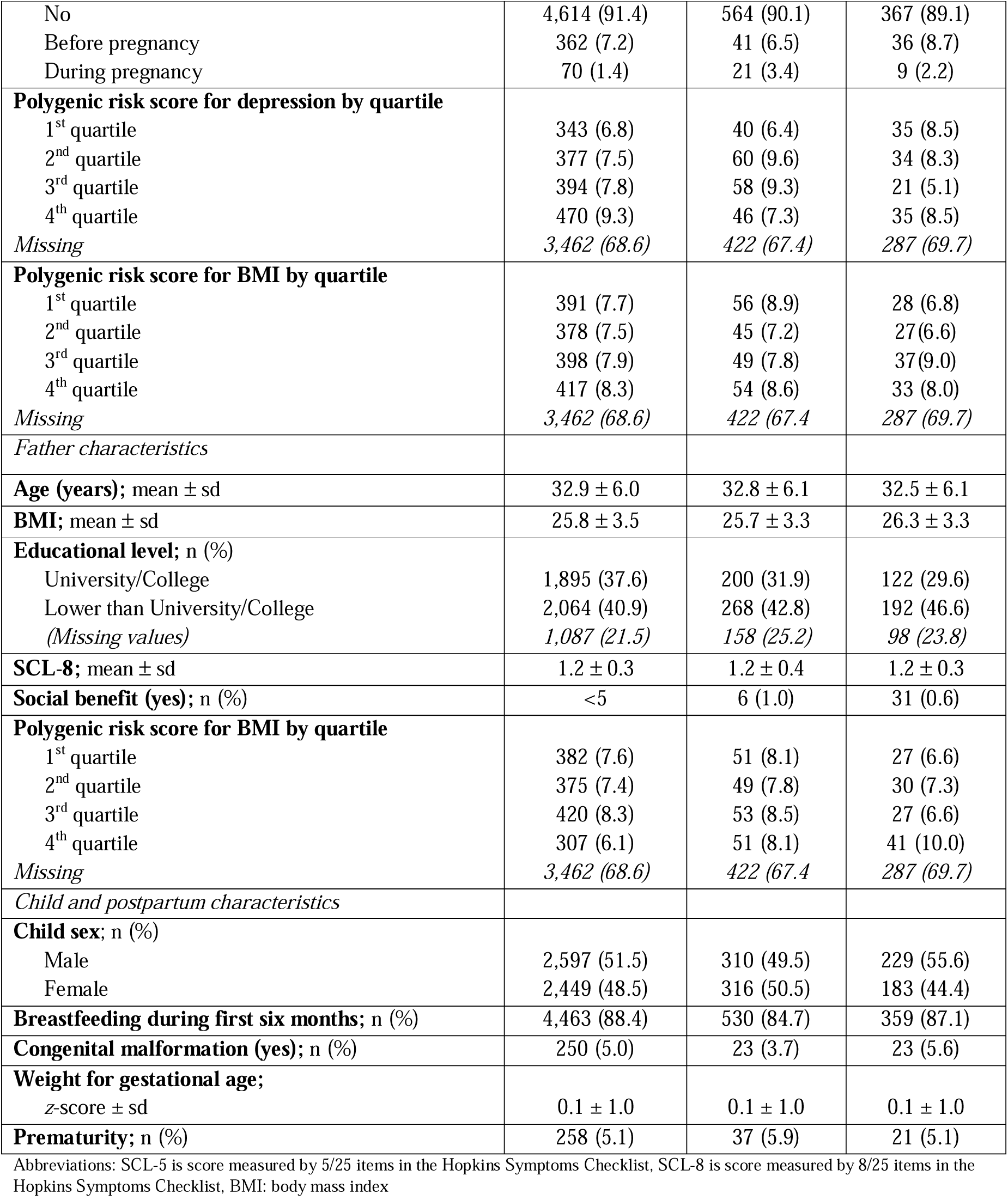
General characteristics of study population according to antidepressant (AD) exposure groups.

### Differences in BMI up to 8 years of age

Figure 2 showed differences in BMI over time between children born to mothers who *continued* antidepressant treatment during pregnancy versus *unexposed* and *discontinued* proximal to pregnancy.

**Figure 2.**
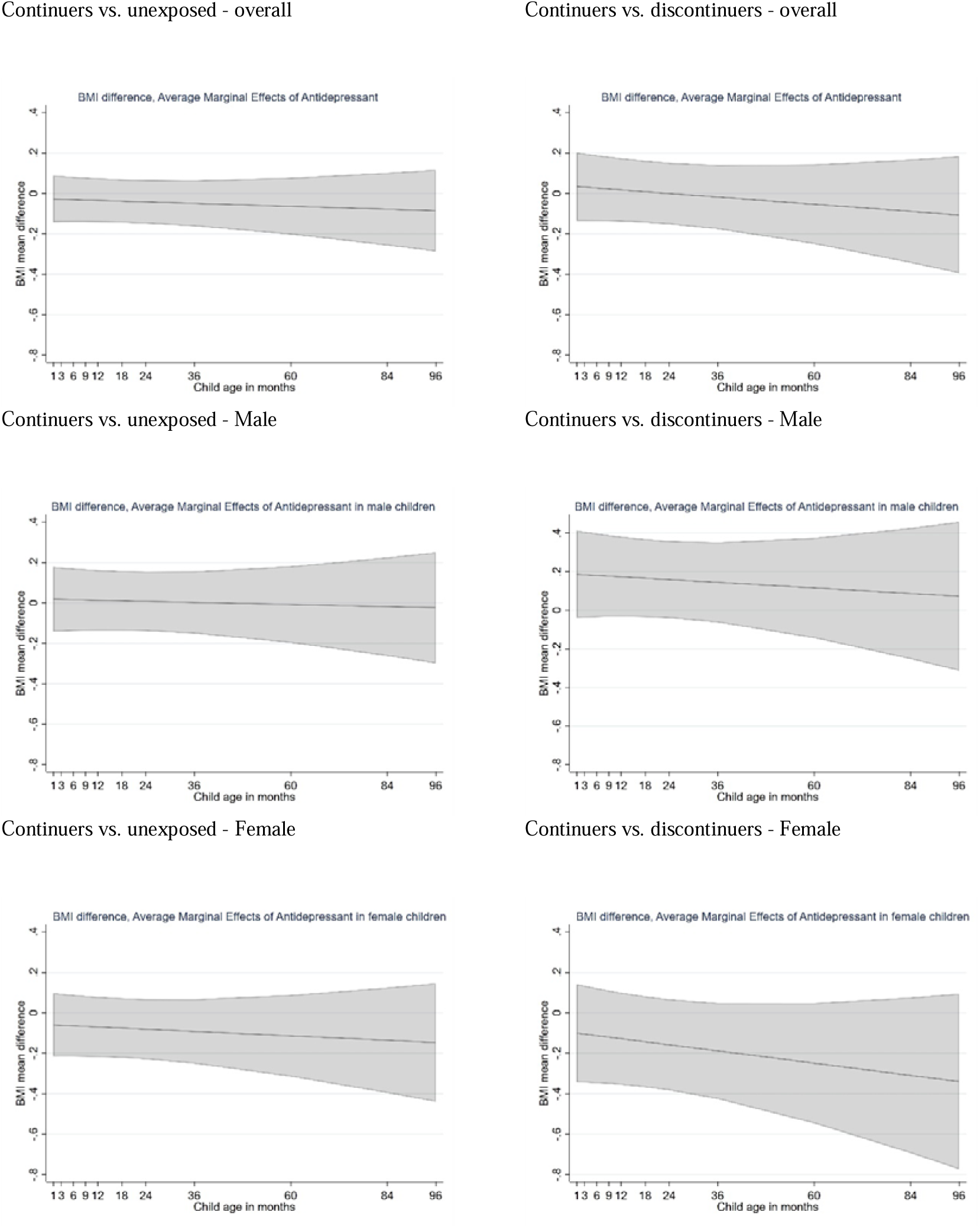
Adjusted mean (±95% CI) differences in body mass index between children born to mothers with pre-pregnancy depression/anxiety who reported using antidepressant before and during pregnancy (continuers) vs. those born to mothers who were unexposed to antidepressant before and during pregnancy (unexposed) or vs. those born to mothers who reported using antidepressant before pregnancy only (discontinuers).

Overall, children born to *continued antidepressant* and *unexposed* mothers had comparable BMI over time (overall mean difference in BMI β= -0.034, 95% CI: -0.149 to 0.081). No significant difference in longitudinal BMI (see Figure 2 and **Table S2**) was seen for both male (β= 0.016, 95% CI: -0.145 to 0.176) and female (β= -0.069, 95% CI: -0.226 to 0.088) offspring.

Children born to *continued antidepressant mothers* also had similar BMI with those born to *discontinued mothers* (β=0.035; 95% CI: -0.134 to 0.204). Slightly higher BMI was observed among male offspring born to *continued antidepressant others* compared to those born to *discontinuers* at baseline (i.e., 6 weeks of age), and the gap became narrower as children grew older. In contrast, lower BMI was seen among female offspring of *continued antidepressant mothers* vs *discontinuers* at baseline and the gap became larger over time (see Figure 2 and **Table S2**).

Children born to mothers with *sustained use of antidepressant* during pregnancy had comparable BMI compared to those born to *antidepressant discontinuers during pregnancy*. Those born to mothers with *low-moderate use of antidepressant* had slightly lower BMI at baseline but higher BMI at 8 years of age compared to children born to *discontinuers during pregnancy*. This pattern was driven by female offspring (β=-0.357; 95% CI: -0.705 to -0.009 at baseline and β=0.343; 95% CI: -0.306 to 0.992 at 8 years). BMI in male offspring born to mothers with *low-moderate antidepressant use* during pregnancy was slightly higher than those born to *discontinuers during pregnancy* throughout their childhood (see Figure 3 and **Table S3**).

**Figure 3.**
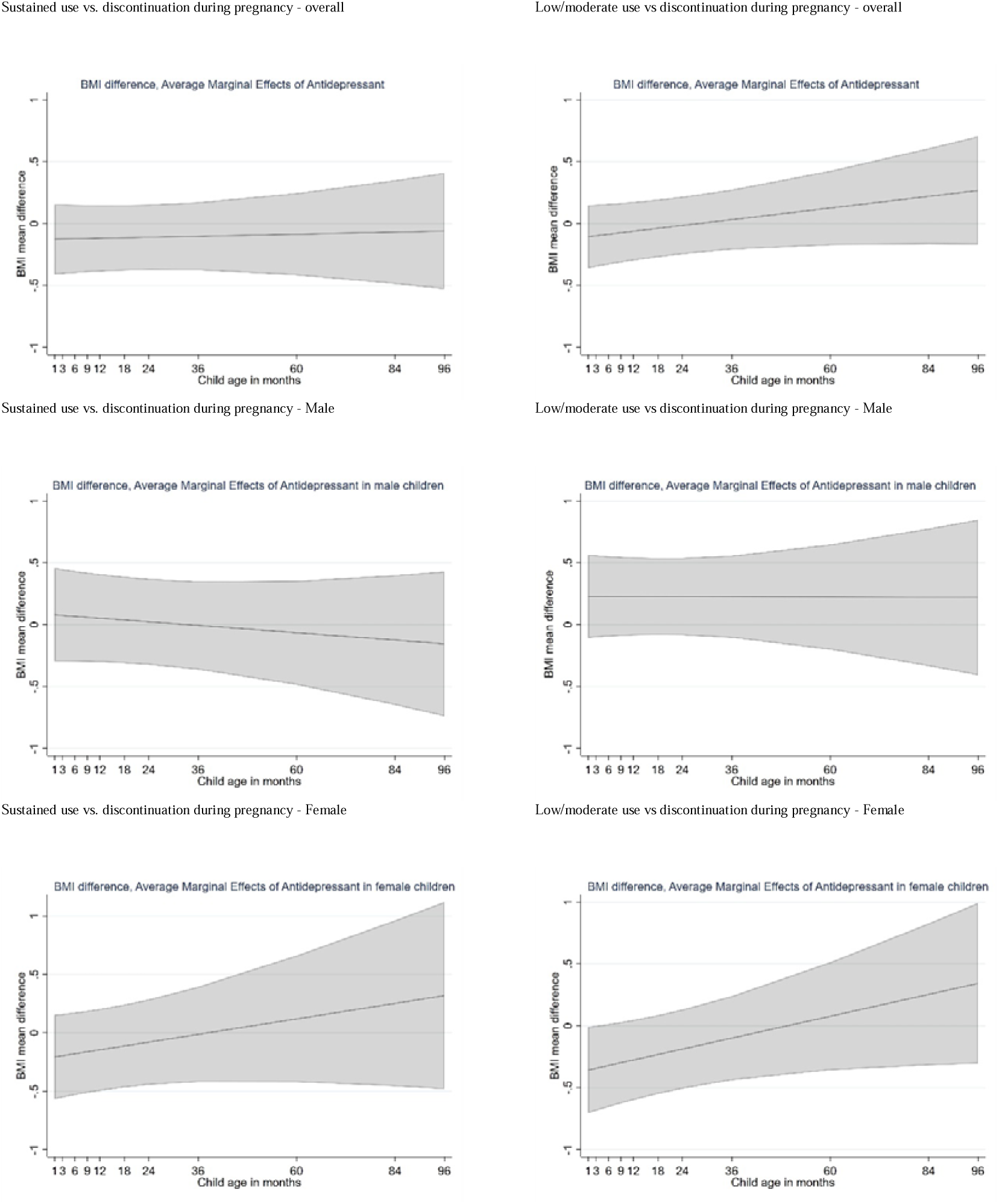
Adjusted mean (±95% CI) differences in body mass index between children born to mothers with pre-pregnancy depression/anxiety with *sustained use of antidepressant during pregnancy* or with *low/moderate use during pregnancy* vs those born to mothers who *discontinued antidepressant during pregnancy*

### Additional analyses

Analyses restricted to children born to SSRI users before pregnancy reveal significantly higher BMI in male children born to *mothers who continued vs discontinued antidepressant* up to 3 years of age (β=0.334; 95% CI: 0.081 to 0.588) and the gap became narrower as the children grew older (see **Table S4**).

Analyses restricted to first pregnancies, term-born children or born with non-low birth weight or taking disease severity into account provided consistent results with main analyses (see **Tables S5-8**).

When main models were additionally adjusted for PRS, we also did not observe significant differences in BMI during childhood across antidepressant exposure trajectories (**Tables S9-10**).

Evaluating BMI changes in children across different antidepressant exposure groups in a subset of parents who had the lowest or highest quartile of PRS for maternal depression, antidepressant response or parental BMI showed divergent results (**Tables S11-18**). In general, we noted potential time- and sex-dependent aspects of the association between antidepressant exposure and childhood BMI in different strata. For example, female children whose mother were in the 1^st^ quartile of PRS (i.e., highest quartile) for depression had lower adjusted BMI in *antidepressant continuers* versus *discontinuers* throughout their childhood (average β= -0.935; 95% CI: -1.844 to -0.227 – see **Table S11**).

## DISCUSSION

This study reports novel findings on the association between antidepressant exposure during pregnancy and longitudinal differences in child BMI from 6 weeks to 8 years of age. In a population of children whose mothers had pre-pregnancy depression/anxiety, those born to mothers who *continued antidepressant during pregnancy* had comparable childhood BMIs with those born to *unexposed mothers*, as well as to mothers who *discontinued treatment proximal to pregnancy.* Similarly, no significant difference in childhood BMI was observed following *sustained prenatal exposure to antidepressant* compared to *discontinuation during pregnancy*. However, we observed lower BMI in female children born to *lower-moderate users* compared to *discontinuers during pregnancy* in early life. In addition, male offspring of *continued antidepressant mothers* had slightly higher BMIs compared to those born to mothers who *discontinued antidepressant proximal to pregnancy* in early life, but the gap faded away as the children grew older. On the other hand, lower BMIs were observed among female offspring born to mothers who *continued antidepressant* vs. *discontinued proximal to pregnancy*, and this difference became larger throughout their childhood. These patterns were particularly stronger and significant in male children born to SSRI users before pregnancy, and in female children born to mothers with *low-moderate use* of antidepressant vs *discontinuation during pregnancy*. Exploratory analyses in a sub-population with genetic data suggest that parental genetic liability for depression and BMI potentially influences these associations but differently in male and female offspring and the impact varies across childhood.

### Interpretations

Although alterations in child growth following prenatal exposure to antidepressants are biologically plausible, few studies have investigated long-term growth outcomes among children prenatally exposed to antidepressants.^8^ In this study, we were able to follow-up the development of the children up to 8 years of age, before the growth spurt at puberty.^29^ Although the general findings point to no differences in childhood BMI across antidepressant exposure groups, the effect size and direction of the association were different between male and female offspring, especially when comparing those born to mothers who *continued antidepressants during pregnancy* versus those who *discontinued the treatment prior to pregnancy*, which is the least confounded comparison.^30^ The underlying mechanism of sex-specific association between prenatal exposure to antidepressant/maternal stress and child growth is supported by a wide range of in vivo studies and consistent with existing human studies.^8,10–12^ A key finding is the lower BMI difference in the first months of life in female children born to mothers with *low-moderate use* of antidepressant, but not with *sustained exposure*. Uncontrolled maternal mental illness during pregnancy due to insufficient antidepressant treatment may well act as dual in-utero exposure and increase the risk of health outcomes and maternal well-being in the early postpartum.^4^ Multiple studies have shown elevated risk for low-birth-weight infant outcomes following prenatal exposure to antidepressant or to maternal psychiatric illness, but co-exposure effects are not yet elucidated.^31,32^ Because low birth weight infants more often display accelerated growth in early life, which poses substantial risks for later central obesity and insulin resistance, our sex-specific results, albeit of moderate effect size, need replicated and further in-depth investigation. Differences in childhood BMI were stronger among male children born to mothers who were SSRI users, with significantly higher BMIs in those born to mothers who *continued treatment into pregnancy* vs *discontinued*. In absolute term, this positive difference is of moderate effect size (about 0.3 difference in BMI) with an upper bound of the 95% CI of 0.6. This finding further supports the biological plausibility of the influence of serotonergic system alterations on child growth,^8^ and partly aligns with prior data for overweight risk in male children at age 7 years.^11^

Maternal polygenic liability for BMI, as evaluated in the lowest and highest quartiles of PRS, showed effect modification influences on our associations, and this was noticeable in the early years of offspring life. Overall, changes in BMI difference in both male and female offspring exhibit temporal patterns throughout childhood, suggesting potential growth reduction, although small, in offspring born to mothers with *antidepressant continuation in pregnancy* compared to those who *discontinued*. This finding highlights the necessity for future studies to investigate child BMI at multiple time points during childhood. Parental metabolic and mental health profiles may influence those in the offspring through shared genetic and environment components. As such, genetic predisposition to mental health conditions and/or elevated BMI inherited from parents might also modify behavioural responses to the environment (e.g., dietary components, physical activity) which further alters the strength of association between genetic component and BMI in the offspring.^28^ In our study, the association between prenatal antidepressant continuation in pregnancy and longitudinal BMI throughout childhood in the offspring differed both based on the effect size and direction of association between those born to parents of higher and lower quartiles of PRSs for depression and BMI. These differences are also sex-dependent, which is in line with previous finding on sex-specific differences in the genetic component of BMI.^33^ Female children, who were born to mothers in the lowest quartile of PRS for depression, had lower adjusted BMI for antidepressant continuers versus discontinuers but this trend was not observed for female children who were born to mothers in the highest quartile of PRS for depression. At early time points after birth, male children who were born to mothers in the lowest and highest quartiles of PRS for BMI had, respectively, lower and higher adjusted BMI for *antidepressant continuers* versus *discontinuers*. Male children whose fathers were in the lowest quartile of PRS for BMI, had lower adjusted BMI in early life for *continuers* versus *unexposed or discontinued* mothers. Taken together, these results suggest the important interplay of maternal predisposition to depression, parental predisposition to BMI, and antidepressant treatment levels during pregnancy in shaping sex-specific growth outcomes in offspring. Further research in independent cohorts is needed to identify the strata of offspring whose growth (measured by change in BMI) is less affected by mothers’ antidepressant continuation into pregnancy.

### Strengths and limitations

This study has several strengths. First, data in MoBa study was collected prospectively which limits potential recall/selection bias after the outcome was known to women. Second, we accounted for maternal depressive/anxiety disorders and measured their symptom severity at different time points in pregnancy using validated instruments. Restriction to women with depression/anxiety at baseline also limited confounding by indication and other factors correlated with maternal illnesses. Third, we addressed attrition (i.e., differential study drop-out) by including all cases with data at one or more of the 11 outcome time-points where BMI was reported. Confounding variability across exposure groups was well-balanced with IPTW method. Fourth, study findings remained robust across several study design specifications, with and without genetic data.

This study has some limitations. First, maternal depressive/anxiety disorders were self-reported by the mother leading to potential reporting bias due to maternal perception about the disease. Second, due to low sample size, we could not examine the effect of antidepressant initiation during pregnancy on child growth. Third, antidepressant use was self-reported leading to potential exposure misclassification bias. However, exposure misclassification of medications for chronic use in MoBa study is low.^34,35^ Fourth, information regarding dosage and duration of antidepressant use is not available in MoBa which precludes further exploration on dose-dependent response. Fifth, outcome measures of child development were self-reported by the mothers which may lead to potential recall bias. Sixth, MoBa study has low response rate with potential selection bias among healthy mothers. Seventh, due to low sample size of children born preterm or with low birth weight, we were not able to perform stratified analyses in these children and thus could not directly examine the role of prematurity and low birth weight on the association of interest. Eighth, our research question focused on prenatal antidepressant use thus we did not examine the impact of antidepressant use and maternal mental health in postpartum period on child growth. Finally, although we took into consideration different study design specifications, residual confounding by depression severity, genetic, environmental, or familial factors cannot be ruled out.

### Conclusion

Longitudinal BMI throughout childhood of children born to mothers with pre-pregnancy depression/anxiety does not differ across prenatal antidepressant exposure trajectories. However, exploratory analyses revealed differences at specific times of growth which might be sex-specific and potentially influenced by parental genetic liability profiles. These findings, if replicated in future research using independent cohorts with larger sample sizes, can inform treatment decision when evaluating risks versus benefits of antidepressant continuation in pregnancy.

## DATA AVAILABILITY STATEMENT

Due to Norwegian legislation, the researcher cannot make the data material on an individual level available to others at any time, including in connection with publication. According to the regulations in Norway and the National Institute of Public Health, no MoBa data on an individual or aggregated level may be uploaded into any repository or database at any time. Data from the Norwegian Mother, Father and Child Cohort Study and the Medical Birth Registry of Norway used in this study are managed by the national health register holders in Norway (Norwegian Institute of Public Health) and can be made available to researchers, provided approval from the Regional Committees for Medical and Health Research Ethics (REC), compliance with the EU General Data Protection Regulation (GDPR) and approval from the data owners. The consent given by the participants does not open for storage of data on an individual level in repositories or journals. Researchers who want access to data sets for replication should apply through helsedata.no.

## ACKNKOWLEDGEMENT

This work, Nhung Trinh and Angela Lupattelli are supported by the Norwegian Research Council (grant no. 288696).

The Norwegian Mother, Father and Child Cohort Study is supported by the Norwegian Ministry of Health and Care Services and the Ministry of Education and Research. We are grateful to all the participating families in Norway who take part in this on-going cohort study. We thank the Norwegian Institute of Public Health (NIPH) for generating high-quality genomic data. This research is part of the HARVEST collaboration, supported by the Research Council of Norway (#229624). We also thank the NORMENT Centre for providing genotype data, funded by the Research Council of Norway (#223273), Southeast Norway Health Authorities and Stiftelsen Kristian Gerhard Jebsen. We further thank the Center for Diabetes Research, the University of Bergen for providing genotype data and performing quality control and imputation of the data funded by the ERC AdG project SELECTionPREDISPOSED, Stiftelsen Kristian Gerhard Jebsen, Trond Mohn Foundation, the Research Council of Norway, the Novo Nordisk Foundation, the University of Bergen, and the Western Norway Health Authorities.

## Notes

### Competing Interest Statement

The authors have declared no competing interest.

### Author Declarations

The current study was approved by The Regional Committees for Medical and Health Research Ethics on 12th February 2020 (reference number: 63566/REK South East), and by the Norwegian Data protection agency (reference number: 672954).

