## Appendix 1 for "Prenatal antidepressant exposure and longitudinal differences in body mass index up to 8 years of age in the offspring born to mothers with pre-pregnancy depressive and/or anxiety in the Norwegian Mother, Father and Child Cohort Study"

**APPENDICES**

**Appendix 1.** Polygenic risk scores

**Figure S1**: Standardized mean difference in confounding factors before and after inverse probability of treatment weighting (IPTW) in children born to mothers who continued antidepressant during pregnancy vs unexposed mothers

**Figure S2**: Standardized mean difference in confounding factors before and after inverse probability of treatment weighting (IPTW) in children born to mothers who continued antidepressant during pregnancy vs those who discontinued antidepressant proximal to pregnancy

**Table S1**. General characteristics of study population according to antidepressant exposure groups during pregnancy

**Table S2**. Weighted differences in children’s BMI (in kg/m2) up to 8 years of age between prenatal antidepressant exposure groups (n=6,084 mother-child pairs), overall and by child sex

**Table S3**. Weighted differences in children’s BMI (in kg/m2) up to 8 years of age between prenatal antidepressant exposure groups (n=426 mother-child pairs), overall and by child sex

**Table S4**. Weighted differences in children’s BMI (in kg/m2) up to 8 years of age between prenatal antidepressant exposure groups (n=5,916 mother-child pairs restricted to users of SSRIs before or during pregnancy and unexposed), overall and by child sex

**Table S5**. Weighted differences in children’s BMI (in kg/m2) up to 8 years of age between prenatal antidepressant exposure groups (n=5,780 mother-child pairs, restricted to first pregnancy), overall and by child sex

**Table S6**. Weighted differences in children’s BMI (in kg/m2) up to 8 years of age between prenatal antidepressant exposure groups (n=5,768 mother-child pairs restricted to term born children), overall and by child sex

**Table S7**. Weighted differences in children’s BMI (in kg/m2) up to 8 years of age between prenatal antidepressant exposure groups (n=5,918 mother-child pairs in non-low-birth -weight children), overall and by child sex

**Table S8**. Weighted differences in children’s BMI (in kg/m2) up to 8 years of age between prenatal antidepressant exposure groups (n=6,084 mother-child pairs), overall and by child sex with adjustment for disease severity measured by the 5 items in the Hopkins Symptoms Checklist

**Table S9**. Weighted differences in children’s BMI (in kg/m2) up to 8 years of age between prenatal antidepressant exposure groups (n=1,913 mother-child pairs in having genetic data), overall and by child sex with additional adjustment for polygenic risks (continuous)

**Table S10**. Weighted differences in children’s BMI (in kg/m2) up to 8 years of age between prenatal antidepressant exposure groups (n=1,913 mother-child pairs in having genetic data), overall and by child sex with additional adjustment for polygenic risks (z-score)

**Table S11**. Weighted differences in children’s BMI (in kg/m2) up to 8 years of age between prenatal antidepressant exposure groups (n=418 mother-child pairs of those in 1^st^ quartile of polygenic risk score for maternal depression), overall and by child sex

**Table S12**. Weighted differences in children’s BMI (in kg/m2) up to 8 years of age between prenatal antidepressant exposure groups (n=551 mother-child pairs in those in 4^th^ quartile of polygenic risk score for maternal depression), overall and by child sex

**Table S13**. Weighted differences in children’s BMI (in kg/m2) up to 8 years of age between prenatal antidepressant exposure groups (n=475 mother-child pairs in those in 1^st^ quartile of polygenic score for maternal BMI), overall and by child sex

**Table S14**. Weighted differences in children’s BMI (in kg/m2) up to 8 years of age between prenatal antidepressant exposure groups (n=504 mother-child pairs in those in 4^th^ quartile of polygenic score for maternal BMI), overall and by child sex

**Table S15**. Weighted differences in children’s BMI (in kg/m2) up to 8 years of age between prenatal antidepressant exposure groups (n=460 mother-child pairs in those in 1^st^ quartile of polygenic score for paternal BMI), overall and by child sex

**Table S16**. Weighted differences in children’s BMI (in kg/m2) up to 8 years of age between prenatal antidepressant exposure groups (n=499 mother-child pairs in those in 4^th^ quartile of polygenic score for paternal BMI), overall and by child sex

**Table S15**. Weighted differences in children’s BMI (in kg/m2) up to 8 years of age between prenatal antidepressant exposure groups (n=460 mother-child pairs in those in 1^st^ quartile of polygenic score for paternal BMI), overall and by child sex

**Table S16**. Weighted differences in children’s BMI (in kg/m2) up to 8 years of age between prenatal antidepressant exposure groups (n=499 mother-child pairs in those in 4^th^ quartile of polygenic score for paternal BMI), overall and by child sex

**Table S17**. Weighted differences in children’s BMI (in kg/m2) up to 8 years of age between prenatal antidepressant exposure groups (n=476 mother-child pairs in those in 1^st^ quartile of polygenic score for maternal antidepressant response), overall and by child sex

**Table S18**. Weighted differences in children’s BMI (in kg/m2) up to 8 years of age between prenatal antidepressant exposure groups (n=480 mother-child pairs in those in 4^th^ quartile of polygenic score for maternal antidepressant response), overall and by child sex

**Appendix 1.** Polygenic risk scores (PRS)

**Description of phenotypes for PRS fitting**

Maternal depression was defined as self-reported depression before and/or during pregnancy (MoBa questionnaires 1 and 3). In these questionnaires, participants were presented with a list of illnesses and asked to report whether they have experienced/were experiencing it and their timing. We used maternal self-reported depression as relevant phenotype because the proportion of this phenotype in MoBa (5-7%) equals estimates of major depression at the time around pregnancy based on based on structural clinical interviews.^1^

Antidepressant response (responders vs non-responders) was defined as 50% reduction of depressive and anxiety smptoms severity, based on the short version (5 items) of the Hopkins Symptoms Checklist (SCL-5) in gestational week 30 (MoBa questionnaire 3) relative to baseline at week 17 (MoBa questionnaire 1).

Maternal Body Mass Index (BMI) was measured before pregnancy started, to avoid overestimation due to gestational weight gain. The pre-pregnancy BMI was based on maternal self-report of weight and height (MoBa questionnaire 1) and calculated as weight in Kg divided by height in meters (m) squared.

**Calculation of PRS**

PRSice version 2.3.3 was used for calculating PRS using the clumping and thresholding (C + T) method.^2,3^ PRS was calculated based on summary statistics from recent genome-wide association studies in European populations for depression and BMI.^4,5^

Effect sizes of single nucleotide polymorphisms (SNP) associations with the phenotype was used to calculate polygenic risk scores (PRS), representing each person’s carriage of all risk variants for the phenotype. Clumping of SNPs, to account for linkage disequilibrium (LD), was performed with the following specifications: clump-kb (the distance for clumping) 250kb, clump-p (the p-value threshold for clumping) 1.0, and clump-r2 (the r2 threshold for clumping) 0.1.

Selection of SNPs for fitting the best PRS for depression in mothers, BMI in mothers, and BMI in fathers

Selection of SNPs was based on the “best-fit” PRS, explaining the highest phenotypic variance, for a selection of participants where we had both the genotype and phenotype data available (5354 for depression, 5526 for BMI in mothers, and 2237 for BMI in fathers). The “best-fit” PRS was calculated at a range of GWAS p-value thresholds starting from 5e-08 (generally regarded as “genome-wide significant”), at 5e-05 intervals. For depression in mothers, all clumped (GWAS p-value threshold 1) SNPs were included (221,739 SNPs) (thereafter called “maternal depression PRS”). For BMI in mother, 29,414 clumped SNPs (GWAS p-value threshold 0.033, (thereafter called “maternal BMI PRS”), and for BMI in father, 50,318 clumped SNPs (GWAS p-value threshold 0.13) were included (thereafter called “paternal BMI PRS”).

The following bar plot shows model fit of the PRS for depression at different P-value thresholds from base GWAS for depression. The best-fitting PRS for depression, explaining the highest phenotypic variance, included all (GWAS p-value threshold 1) clumped SNPs (221,739 SNP).

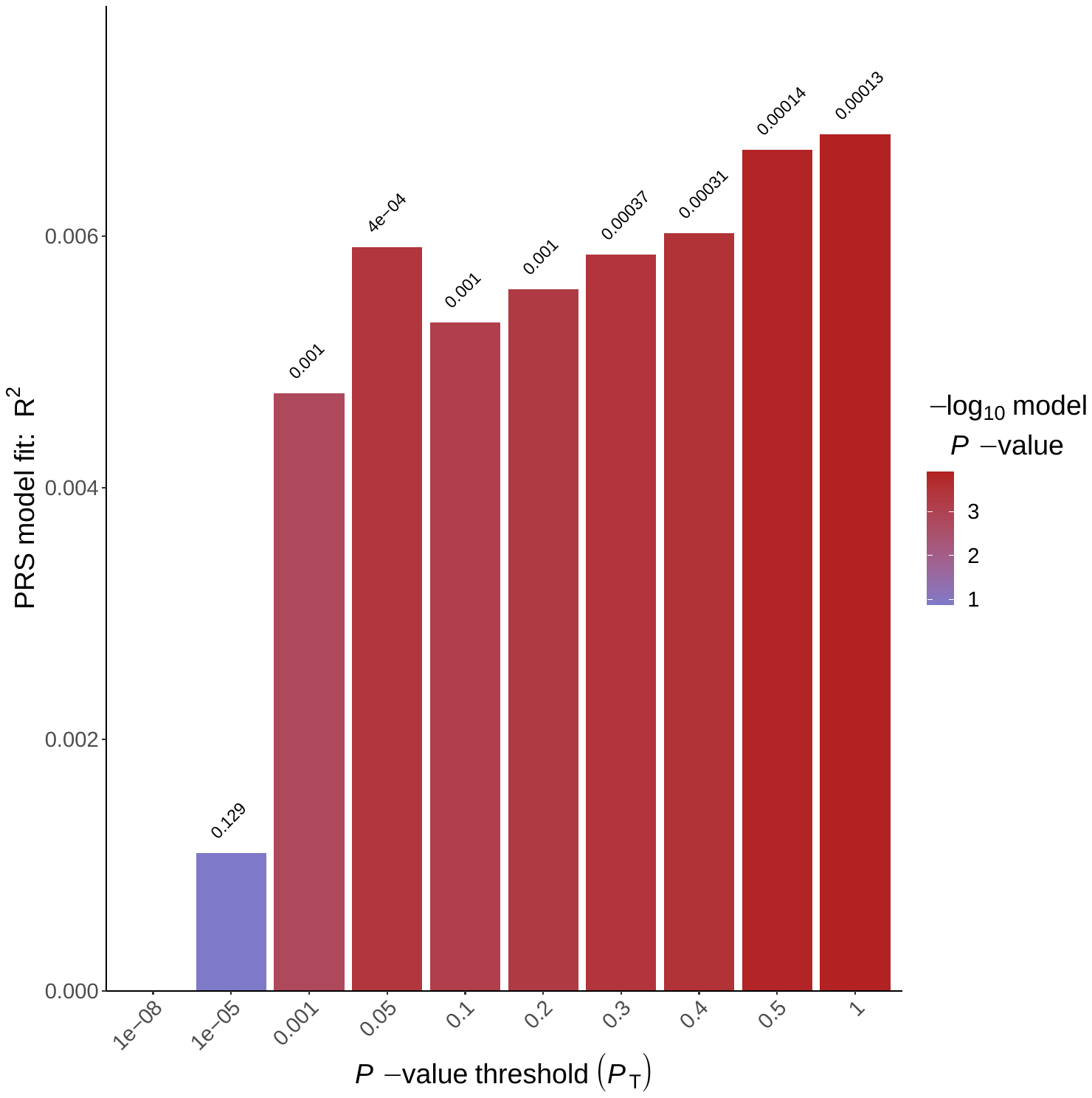

The following quantile plot (with 20 evenly spaced quantiles), based on the best-fitting PRS for depression, shows the effect of increasing PRS on predicted risk of depression. Note that the middle quantile (quantile 10) was used as the reference.

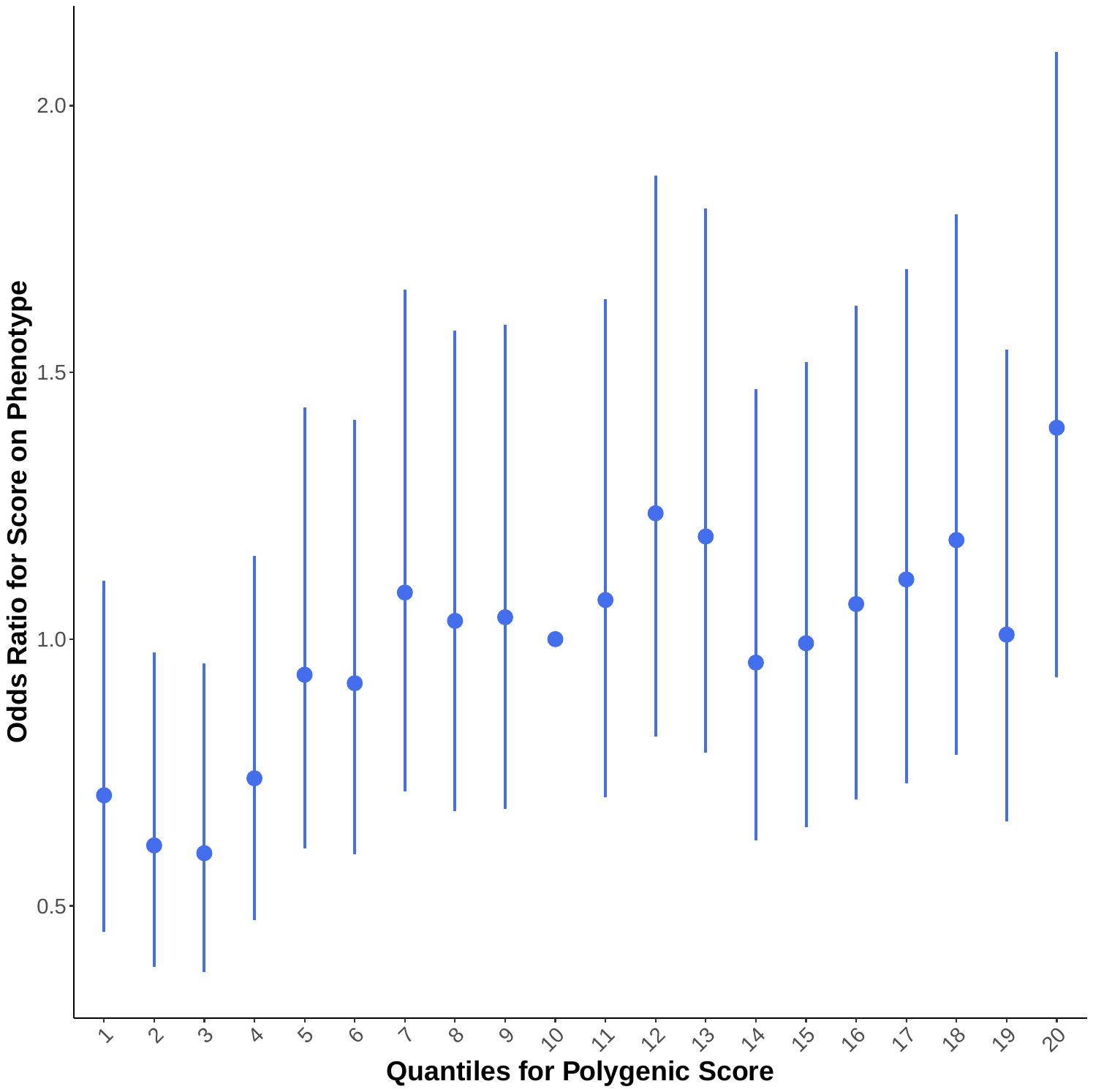

The following bar plot shows model fit of the PRS for BMI in mothers at different P-value thresholds from base GWAS for BMI. The best-fitting PRS for BMI in mothers, explaining the highest phenotypic variance, was at GWAS p-value threshold of 0.033 and included 29,414 clumped SNPs.

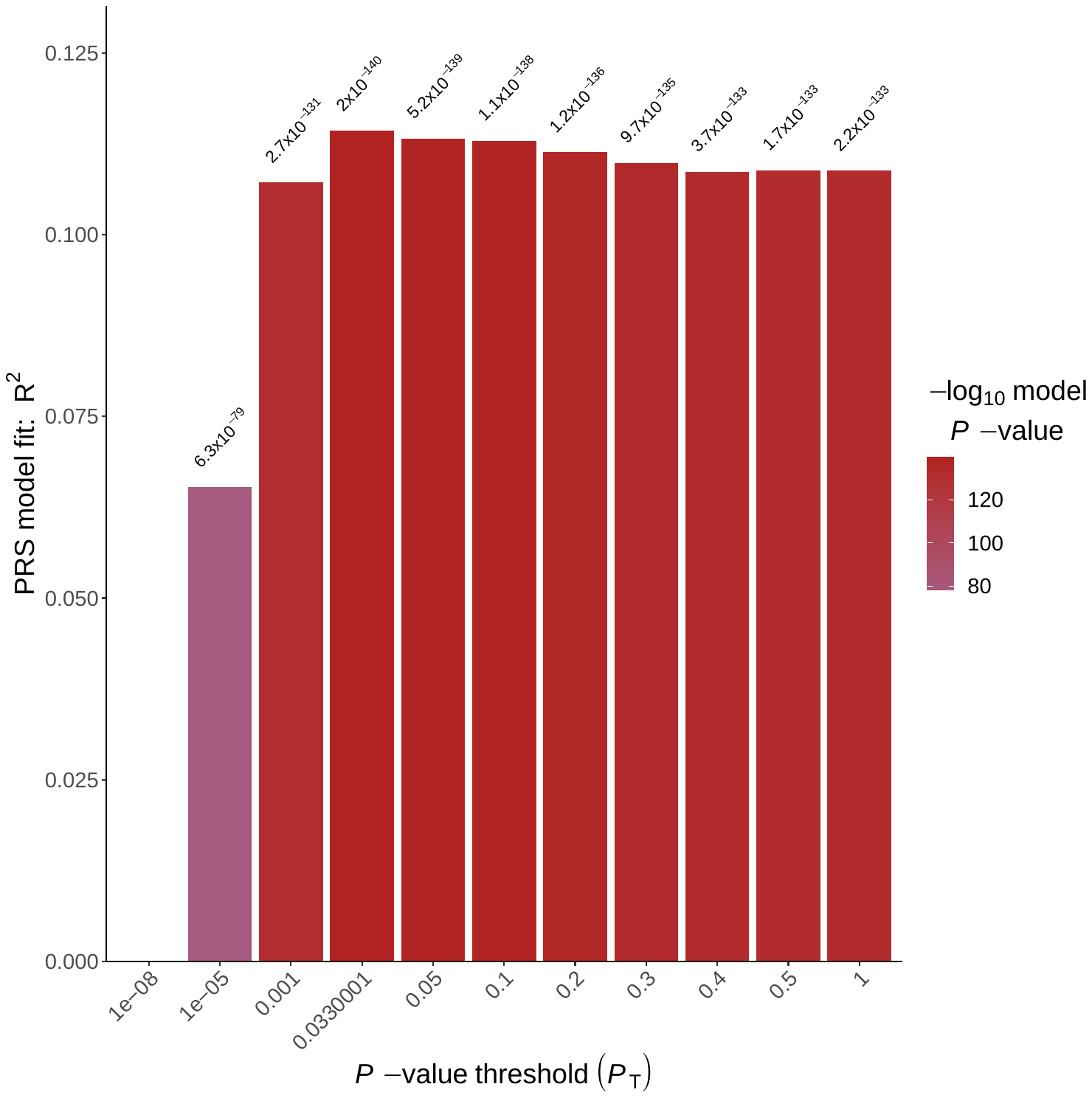

The following quantile plot (with 20 evenly spaced quantiles), based on the best-fitting PRS for BMI in mothers, show the effect of increasing PRS on change in BMI of mothers. Note that the middle quantile (quantile 10) was used as the reference.

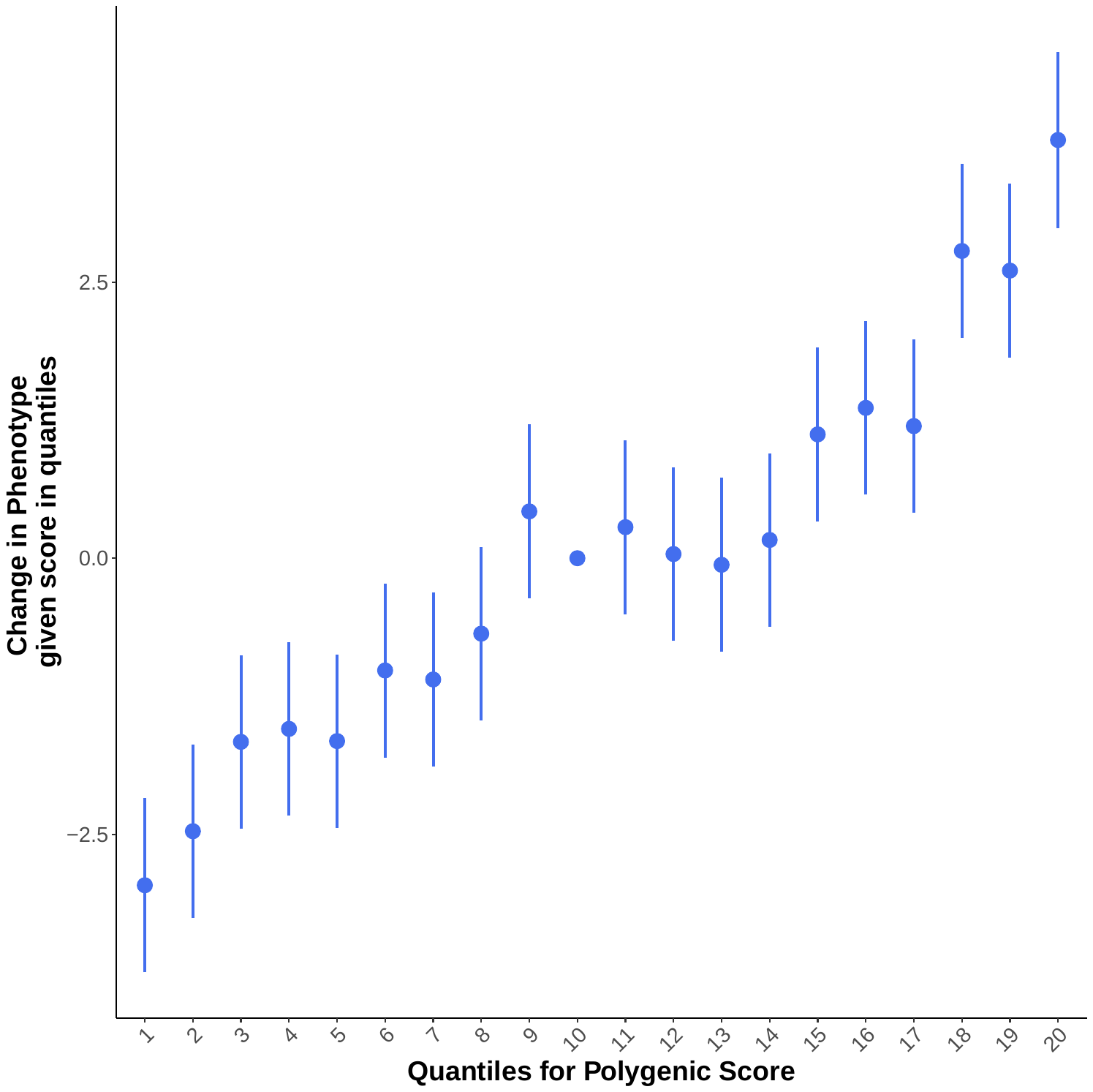

The following bar plot shows model fit of the PRS for BMI in fathers at different P-value thresholds from base GWAS for BMI. The best-fitting PRS for BMI in fathers, explaining the highest phenotypic variance, was at GWAS p-value threshold of 0.13 and included 50,318 clumped SNPs.

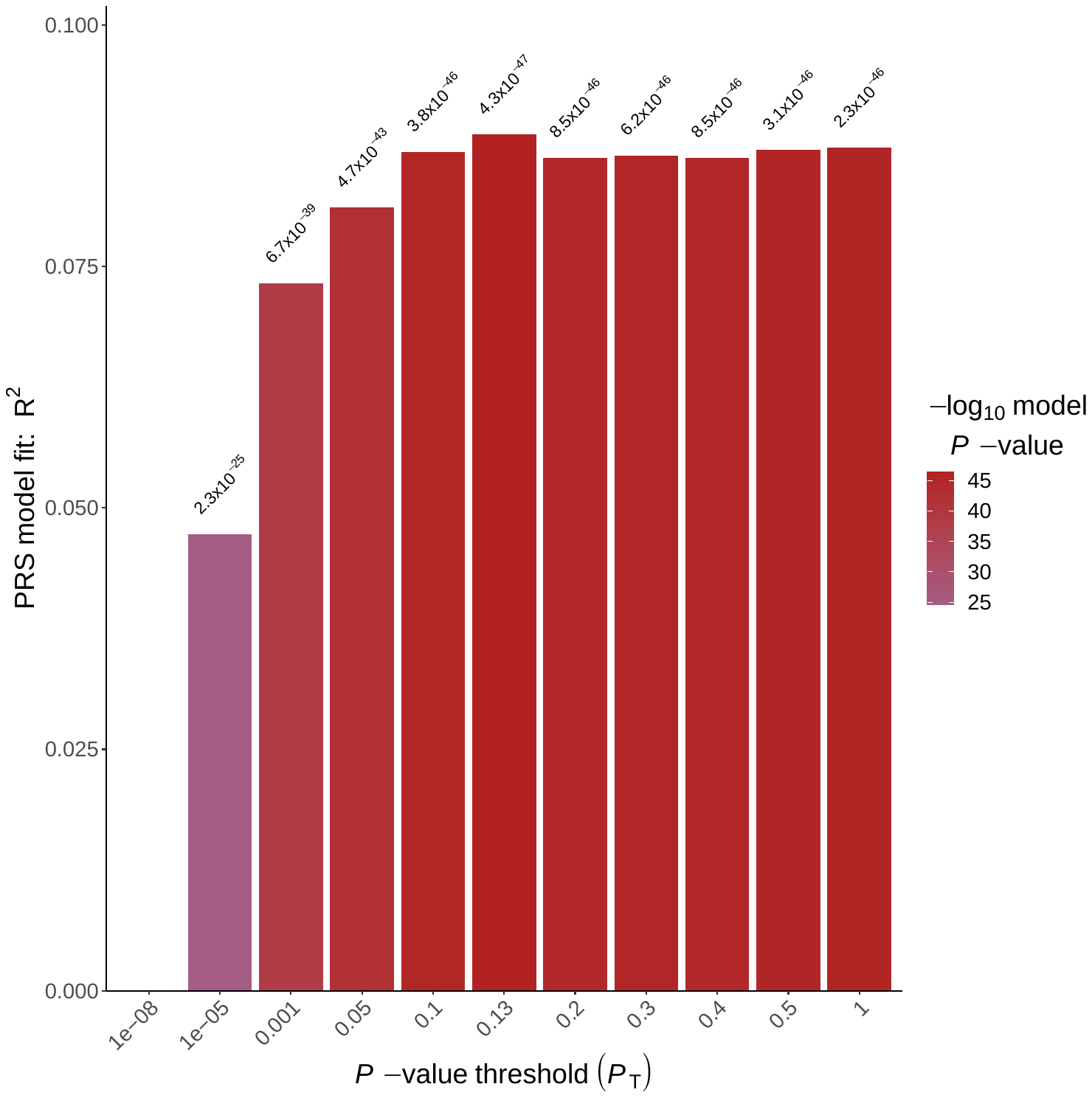

The following quantile plot (with 20 evenly spaced quantiles), based on the best-fitting PRS for BMI in fathers, show the effect of increasing PRS on change in BMI of fathers. Note that the middle quantile (quantile 10) was used as the reference.

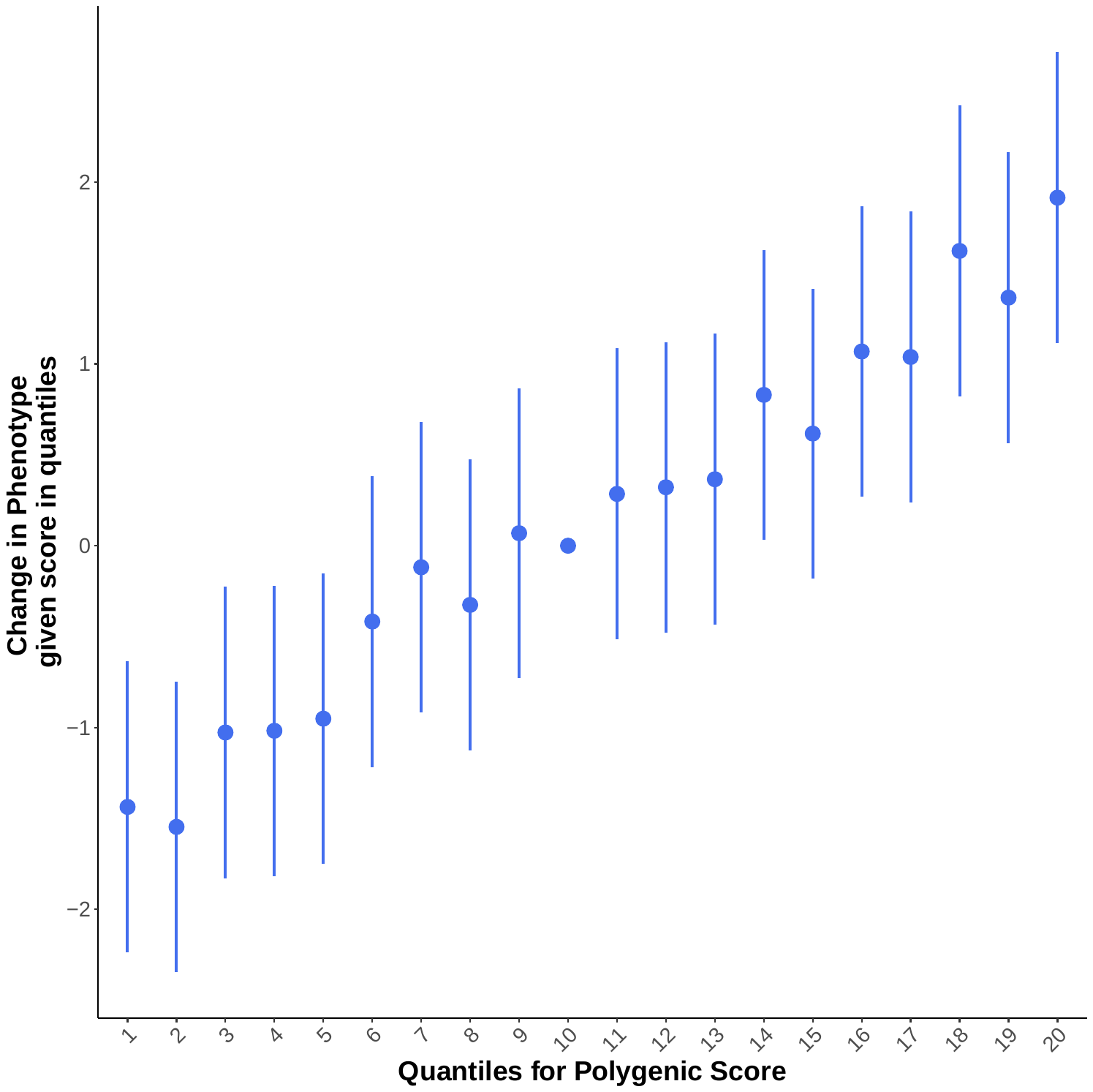

Selection of SNPs for anti-depressant response

To our knowledge, there is currently not many published GWAS with large sample sizes for anti-depressant response. We used non-remission antidepressant response GWAS summary statistics for European population from Pain et al study, and rather than using the GWAS p-value thresholding method, we only used SNPs with GWAS p-value <5.005e-05.^6^ Our PRS for antidepressant response was based on 27 SNPs.

**Figure S1**: Standardized mean difference in confounding factors before and after inverse probability of treatment weighting in children born to mothers who continued antidepressant during pregnancy vs unexposed mothers

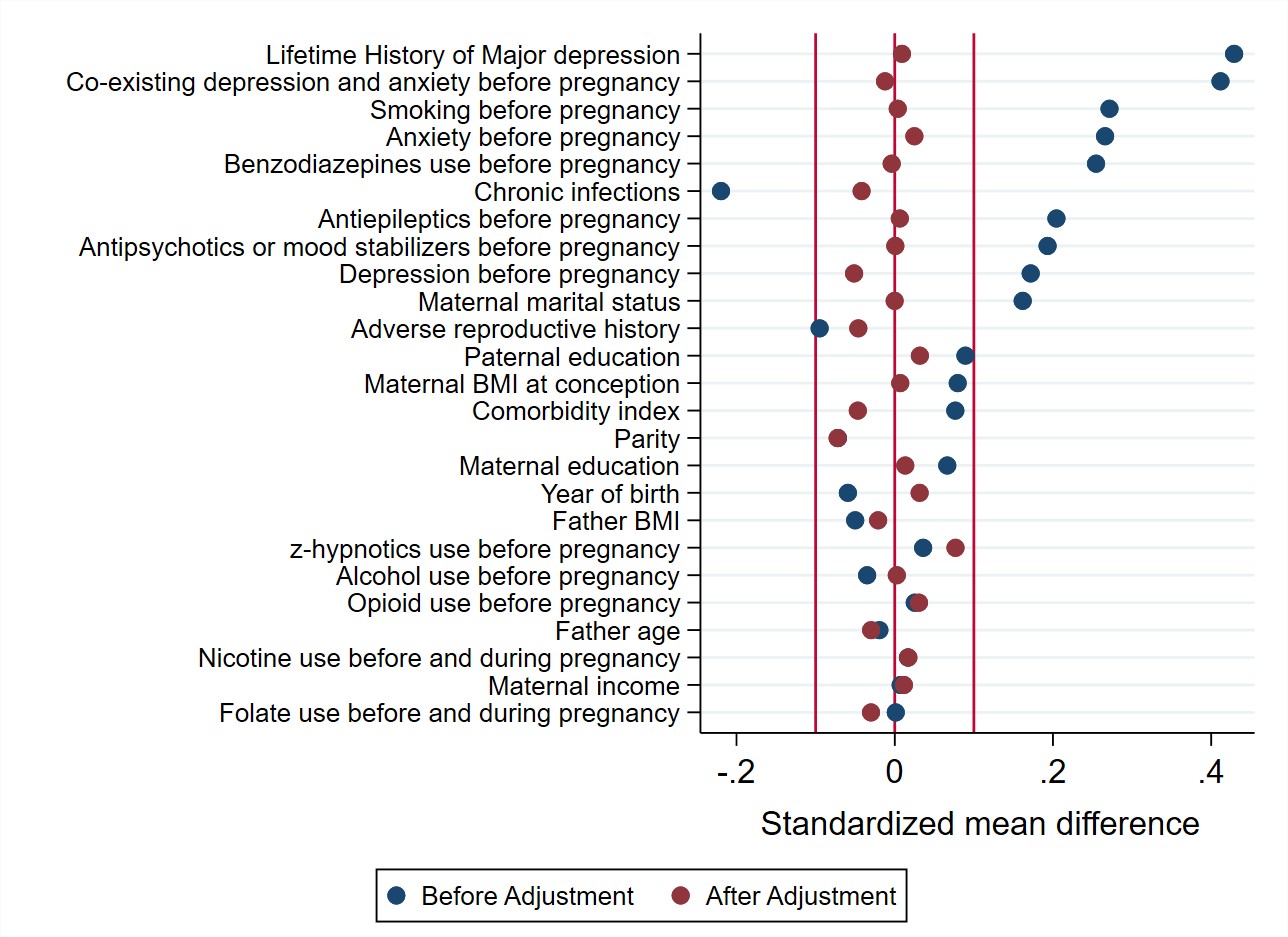

**Figure S2**: Standardized mean difference in confounding factors before and after inverse probability of treatment weighting in children born to mothers who continued antidepressant during pregnancy vs those who discontinued antidepressant proximal to pregnancy

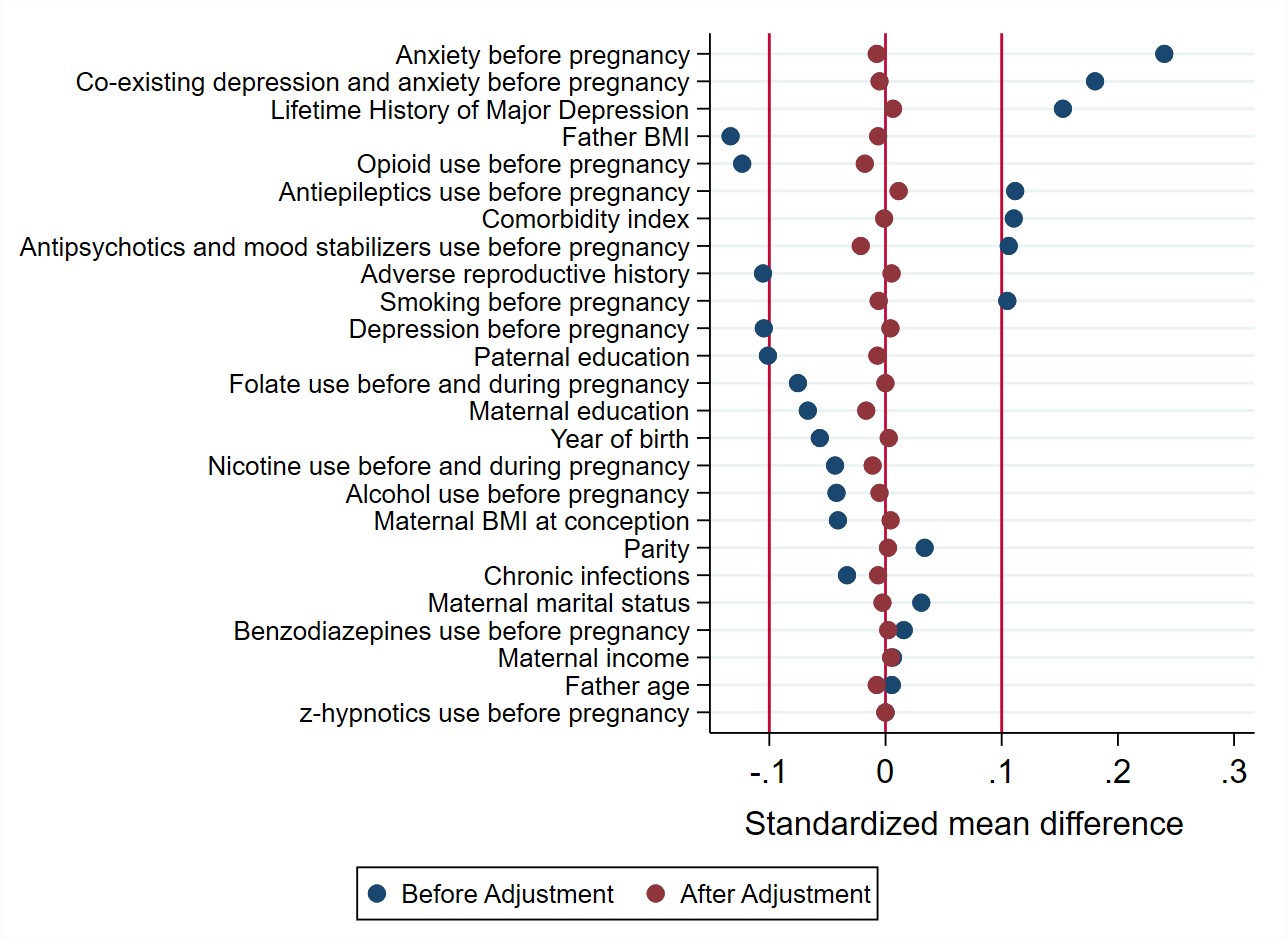

**Table S1.** General characteristics of study population according to antidepressant exposure groups during pregnancy

| Characteristics | Sustained use | Low/moderate use | Discontinuation during pregnancy |
| --- | --- | --- | --- |
| *Mother characteristics* |  |  |  |
| **Age (years);** mean ± sd | 31.0 ± 5.1 | 2.9 ± 4.9 | 30.0 ± 5.1 |
| **Married/cohabiting;** n (%) | 159 (91.9) | 187 (87.8) | 204 (85.0) |
| **Primiparous (yes); n(%)** | 89 (51.4) | 125 (58.7) | 131 (54.6) |
| **Educational level**; n (%)  University/College  Lower than University/College | 104 (60.1)  69 (39.9) | 114 (53.5)  99 (46.5) | 121 (50.4)  119 (49.6) |
| **Grossly yearly income;** n (%)  Average  Low  High  (*Missing values*) | 109 (63.0)  49 (28.3)  12 (6.9)  *<5* | 123 (57.7)  65 (30.5)  20 (9.4)  *5 (2.3)* | 149 (62.1)  66 (27.5)  16 (6.7)  *9 (3.8)* |
| **Obstetric risk score;** mean ± sd | 0.5 ± 0.8 | 0.5 ± 0.8 | 0.4 ± 0.8 |
| **Folate intake before and/or during pregnancy (yes);** n (%) | 105 (60.7) | 130 (61.0) | 143 (59.6) |
| **Smoking up until returning questionnaire 1;** n (%)  No  Yes  Stopped at pregnancy  (*Missing values*) | 111 (64.2)  36 (20.8)  26 (15.0)  - | 116 (54.5)  43 (20.2)  52 (24.4)  *<5* | 135 (56.2)  44 (18.3)  61 (25.4)  - |
| **Comorbid depression and anxiety; n(%)** | 91 (52.6) | 97 (45.5) | 88 (36.7) |
| **Life time history of major depression (yes);** n (%)  *(Missing values)* | 91 (52.6)  *<5* | 110 (51.6)  *<5* | 96 (40.0)  *<5* |
| **SCL-5 at gestational week 17;** mean ± sd | 1.9 ± 0.8 | 1.9 ± 0.6 | 1.8 ± 0.7 |
| **SCL-5 at gestational week 30;** mean ± sd | 1.8 ± 0.7 | 1.9 ± 0.7 | 1.8 ± 0.7 |
| **Co-medications use before pregnancy (yes);** n (%)  Antiepileptic drugs  Antipsychotics  Benzodiazepines  Hypnotics  Opioids | 7 (4.0)  6 (3.5)  16 (9.2)  <5  6 (3.5) | 8 (3.8)  12 (5.6)  15 (7.0)  <5  6 (2.8) | 5 (2.1)  5 (2.1)  22 (9.2)  <5  7 (2.9) |
| **Alcohol use in pregnancy;** n (%)  No/very limited  Medium use  Weekly use  *Missing values* | 143 (82.7)  20 (11.6)  <5  *6 (3.5)* | 178 (83.6)  29 (13.6)  <5  *<5* | 203 (84.6)  28 (11.7)  5 (2.1)  *<5* |
| **Illicit substances use (yes);** n (%) | <5 | 13 (6.1) | 10 (4.2) |
| **Nicotine use (yes);** n (%)  No  Before pregnancy  During pregnancy | 153 (88.4)  12 (6.9)  8 (4.6) | 196 (92.0)  12 (5.6)  5 (2.3) | 215 (89.6)  17 (7.1)  8 (3.3) |
| *Father characteristics* |  |  |  |
| **Age (years);** mean ± sd | 33.3 ± 5.8 | 32.3 ± 6.0 | 33.0 ± 6.3 |
| **BMI;** mean ± sd | 25.4 ± 3.1 | 26.0 ± 3.7 | 25.6 ± 3.0 |
| **Educational level;** n (%)  University/College  Lower than University/College  *(Missing values)* | 56 (32.4)  79 (45.7)  *38 (22.0)* | 81 (38.0)  84 (39.4)  *48 (22.5)* | 63 (26.2)  105 (43.8)  *72 (30.0)* |
| **SCL-8;** mean ± sd | 1.1 ± 0.2 | 1.3 ± 0.5 | 1.2 ± 0.4 |
| **Social benefit (yes);** n (%) | - | <5 | <5 |
| *Child and postpartum characteristics* |  |  |  |
| **Child sex**; n (%)  Male  Female | 91 (52.6)  82 (47.4) | 90 (42.3)  123 (57.7) | 129 (53.8)  111 (46.2) |
| **Breastfeeding during first six months;** n (%) | 141 (81.5) | 182 (85.4) | 207 (86.2) |
| **Congenital malformation (yes);** n (%) | 5 (2.9) | 7 (3.3) | 11 (4.6) |
| **Weight for gestational age;**  *z*-score ± sd | -0.1 ± 0.9 | 0.0 ± 0.9 | 0.2 ± 1.0 |
| **Prematurity;** n (%) | 13 (7.5) | 13 (6.1) | 11 (4.6) |

Abbreviations: SCL-5 is score measured by 5/25 items in the Hopkins Symptoms Checklist, SCL-8 is score measured by 8/25 items in the Hopkins Symptoms Checklist, BMI: body mass index

**Table S2**. Weighted differences in children’s BMI (in kg/m2) up to 8 years of age between prenatal antidepressant exposure groups (n=6,084 mother-child pairs), overall and by child sex

|  | BMI (in kg/m2) | Antidepressant exposure | | | |
| --- | --- | --- | --- | --- | --- |
|  |  | Continuers vs discontinuers | | Continuers vs unexposed | |
|  | Mean (SD) | beta | 95%CI | beta | 95%CI |
| **All children** |  |  |  |  |  |
| Average | 16.52 (1.27) | 0.035 | -0.134, 0.204 | -0.034 | -0.149, 0.081 |
| By age |  |  |  |  |  |
| 1 month | 15.50 (1.63) | 0.034 | -0.134, 0.201 | -0.035 | -0.148, 0.079 |
| 3 months | 16.60 (1.55) | 0.031 | -0.134, 0.196 | -0.036 | -0.148, 0.076 |
| 6 months | 17.21 (1.54) | 0.026 | -0.135, 0.188 | -0.038 | -0.148, 0.072 |
| 8 months | 17.35 (1.51) | 0.022 | -0.136, 0.180 | -0.039 | -0.148, 0.069 |
| 12 months | 17.01 (1.40) | 0.018 | -0.138, 0.173 | -0.041 | -0.148, 0.066 |
| 18 months | 16.76 (1.34) | 0.009 | -0.143, 0.161 | -0.045 | -0.150, 0.061 |
| 2 years | 16.48 (1.42) | 0.000 | -0.151, 0.151 | -0.049 | -0.154, 0.057 |
| 3 years | 16.09 (1.52) | -0.017 | -0.175, 0.140 | -0.056 | -0.167, 0.055 |
| 5 years | 16.23 (1.96) | -0.053 | -0.248, 0.143 | -0.071 | -0.209, 0.068 |
| 7 years | 15.83 (1.86) | -0.088 | -0.342, 0.167 | -0.085 | -0.263, 0.093 |
| 8 years | 15.56 (1.56) | -0.105 | -0.393, 0.183 | -0.093 | -0.293, 0.108 |
| **Male** |  |  |  |  |  |
| Average | 16.70 (1.25) | 0.185 | -0.045, 0.414 | 0.016 | -0.145, 0.176 |
| By age |  |  |  |  |  |
| 1 month | 15.77 (1.68) | 0.183 | -0.044, 0.410 | 0.015 | -0.144, 0.174 |
| 3 months | 16.91 (1.54) | 0.181 | -0.042, 0.404 | 0.014 | -0.142, 0.171 |
| 6 months | 17.42 (1.51) | 0.177 | -0.040, 0.395 | 0.013 | -0.140, 0.166 |
| 8 months | 17.54 (1.49) | 0.174 | -0.039, 0.386 | 0.011 | -0.139, 0.162 |
| 12 months | 17.23 (1.37) | 0.170 | -0.039, 0.379 | 0.010 | -0.138, 0.158 |
| 18 months | 16.95 (1.31) | 0.163 | -0.040, 0.366 | 0.007 | -0.138, 0.153 |
| 2 years | 16.58 (1.36) | 0.155 | -0.046, 0.357 | 0.005 | -0.141, 0.150 |
| 3 years | 16.17 (1.50) | 0.141 | -0.068, 0.350 | -0.001 | -0.153, 0.151 |
| 5 years | 16.19 (1.88) | 0.112 | -0.149, 0.372 | -0.012 | -0.200, 0.176 |
| 7 years | 15.84 (1.78) | 0.083 | -0.259, 0.424 | -0.023 | -0.265, 0.220 |
| 8 years | 15.57 (1.51) | 0.068 | -0.319, 0.455 | -0.028 | -0.302, 0.246 |
| **Female** |  |  |  |  |  |
| Average | 16.33 (1.27) | -0.092 | -0.334, 0.150 | -0.069 | -0.226, 0.088 |
| By age |  |  |  |  |  |
| 1 month | 15.22 (1.53) | -0.094 | -0.334, 0.146 | -0.070 | -0.226, 0.086 |
| 3 months | 16.27 (1.49) | -0.099 | -0.336, 0.137 | -0.072 | -0.226, 0.082 |
| 6 months | 16.98 (1.53) | -0.107 | -0.338, 0.125 | -0.075 | -0.226, 0.076 |
| 8 months | 17.15 (1.50) | -0.114 | -0.342, 0.113 | -0.077 | -0.226, 0.071 |
| 12 months | 16.78 (1.39) | -0.122 | -0.346, 0.103 | -0.080 | -0.227, 0.067 |
| 18 months | 16.55 (1.35) | -0.136 | -0.358, 0.085 | -0.085 | -0.231, 0.060 |
| 2 years | 16.38 (1.49) | -0.151 | -0.373, 0.070 | -0.091 | -0.238, 0.056 |
| 3 years | 15.99 (1.54) | -0.181 | -0.415, 0.052 | -0.102 | -0.259, 0.055 |
| 5 years | 16.27 (2.05) | -0.241 | -0.535, 0.053 | -0.123 | -0.322, 0.076 |
| 7 years | 15.84 (1.95) | -0.301 | -0.683, 0.081 | -0.145 | -0.403, 0.113 |
| 8 years | 15.55 (1.62) | -0.331 | -0.762, 0.100 | -0.156 | -0.447, 0.135 |

All models are weighted for sociodemographic-lifestyle-reproductive factors (i.e., maternal age, parity, marital status, obstetric comorbidity index adapted from Bateman et al., maternal education, maternal gross yearly income, smoking and alcohol use before pregnancy, BMI at conception, history of abortions/miscarriages, folic acid intake, paternal age, paternal education, paternal social benefits, and paternal BMI), maternal psychiatric correlates (i.e., Lifetime History of Major Depression measured in Q1), comedication before pregnancy (i.e., opioid analgesics – ATC code N02A, benzodiazepines/z-hypnotics - ATC codes N05B and N05C, antipsychotics and mood stabilizers - ATC code N05A, and antiepileptics - ATC code N03A) using inverse probability of treatment weighting.

**Table S3**. Weighted differences in children’s BMI (in kg/m2) up to 8 years of age between prenatal antidepressant exposure groups (n=426 mother-child pairs), overall and by child sex

|  | Antidepressant exposure | | | |
| --- | --- | --- | --- | --- |
|  | Sustained use vs discontinuers in pregnancy | | Low-moderate use vs discontinuers in pregnancy | |
|  | beta | 95%CI | beta | 95%CI |
| **All children** |  |  |  |  |
| Average | -0.128 | -0.414, 0.158 | -0.109 | -0.364, 0.147 |
| By age |  |  |  |  |
| 1 month | -0.127 | -0.411, 0.157 | -0.105 | -0.358, 0.149 |
| 3 months | -0.126 | -0.406, 0.155 | -0.097 | -0.347, 0.153 |
| 6 months | -0.123 | -0.399, 0.152 | -0.085 | -0.330, 0.159 |
| 8 months | -0.121 | -0.393, 0.150 | -0.073 | -0.313, 0.167 |
| 12 months | -0.119 | -0.388, 0.149 | -0.062 | -0.298, 0.175 |
| 18 months | -0.115 | -0.380, 0.150 | -0.038 | -0.270, 0.194 |
| 2 years | -0.111 | -0.375, 0.154 | -0.015 | -0.246, 0.217 |
| 3 years | -0.102 | -0.377, 0.173 | 0.033 | -0.210, 0.275 |
| 5 years | -0.085 | -0.417, 0.247 | 0.127 | -0.173, 0.427 |
| 7 years | -0.068 | -0.487, 0.351 | 0.221 | -0.167, 0.608 |
| 8 years | -0.060 | -0.529, 0.409 | 0.268 | -0.169, 0.705 |
| **Male** |  |  |  |  |
| Overall | 0.082 | -0.297, 0.463 | 0.229 | -0.109, 0.568 |
| By age |  |  |  |  |
| 1 month | 0.080 | -0.297, 0.458 | 0.229 | -0.107, 0.565 |
| 3 months | 0.075 | -0.297, 0.448 | 0.229 | -0.102, 0.560 |
| 6 months | 0.068 | -0.298, 0.434 | 0.229 | -0.095, 0.552 |
| 8 months | 0.061 | -0.299, 0.421 | 0.228 | -0.090, 0.547 |
| 12 months | 0.053 | -0.302, 0.409 | 0.228 | -0.086, 0.542 |
| 18 months | 0.038 | -0.310, 0.387 | 0.228 | -0.082, 0.538 |
| 2 years | 0.024 | -0.323, 0.370 | 0.227 | -0.085, 0.539 |
| 3 years | -0.006 | -0.362, 0.350 | 0.226 | -0.106, 0.558 |
| 5 years | -0.065 | -0.485, 0.354 | 0.224 | -0.201, 0.649 |
| 7 years | -0.125 | -0.648, 0.399 | 0.222 | -0.334, 0.777 |
| 8 years | -0.154 | -0.739, 0.431 | 0.221 | -0.407, 0.849 |
| **Female** |  |  |  |  |
| Overall | -0.212 | -0.576, 0.152 | **-0.364** | **-0.716, -0.013** |
| By age |  |  |  |  |
| 1 month | -0.206 | -0.568, 0.156 | **-0.357** | **-0.705, -0.009** |
| 3 months | -0.195 | -0.553, 0.163 | -0.342 | -0.685, 0.002 |
| 6 months | -0.178 | -0.532, 0.175 | -0.320 | -0.655, 0.015 |
| 8 months | -0.162 | -0.513, 0.189 | -0.298 | -0.627, 0.030 |
| 12 months | -0.145 | -0.495, 0.205 | -0.276 | -0.600, 0.048 |
| 18 months | -0.112 | -0.466, 0.242 | -0.232 | -0.550, 0.087 |
| 2 years | -0.079 | -0.445, 0.287 | -0.187 | -0.507, 0.132 |
| 3 years | -0.012 | -0.420, 0.395 | -0.099 | -0.438, 0.240 |
| 5 years | 0.121 | -0.421, 0.662 | 0.078 | -0.358, 0.513 |
| 7 years | 0.254 | -0.457, 0.964 | 0.255 | -0.318, 0.827 |
| 8 years | 0.320 | -0.481, 1.121 | 0.343 | -0.306, 0.992 |

All models are weighted for sociodemographic-lifestyle-reproductive factors (i.e., maternal age, parity, marital status, obstetric comorbidity index adapted from Bateman et al., maternal education, maternal gross yearly income, smoking and alcohol use before pregnancy, BMI at conception, history of abortions/miscarriages, folic acid intake, paternal age, paternal education, paternal social benefits, and paternal BMI), maternal psychiatric correlates (i.e., Lifetime History of Major Depression measured in Q1), comedication before pregnancy (i.e., opioid analgesics – ATC code N02A, benzodiazepines/z-hypnotics - ATC codes N05B and N05C, antipsychotics and mood stabilizers - ATC code N05A, and antiepileptics - ATC code N03A) using inverse probability of treatment weighting.

**Table S4**. Weighted differences in children’s BMI (in kg/m2) up to 8 years of age between prenatal antidepressant exposure groups (n=5,916 mother-child pairs restricted to users of SSRIs before or during pregnancy and unexposed), overall and by child sex

|  | Antidepressant exposure | | | |
| --- | --- | --- | --- | --- |
|  | SSRI continuers vs discontinuers | | SSRI continuers vs unexposed | |
|  | beta | 95%CI | beta | 95%CI |
| **All children** |  |  |  |  |
| Average | 0.144 | -0.046, 0.334 | 0.021 | -0.106, 0.147 |
| By age |  |  |  |  |
| 1 month | 0.142 | -0.046, 0.331 | 0.020 | -0.106, 0.145 |
| 3 months | 0.138 | -0.047, 0.323 | 0.018 | -0.105, 0.142 |
| 6 months | 0.132 | -0.049, 0.313 | 0.016 | -0.105, 0.137 |
| 8 months | 0.125 | -0.052, 0.303 | 0.014 | -0.106, 0.133 |
| 12 months | 0.119 | -0.055, 0.293 | 0.011 | -0.106, 0.129 |
| 18 months | 0.106 | -0.063, 0.276 | 0.007 | -0.109, 0.122 |
| 2 years | 0.094 | -0.074, 0.262 | 0.002 | -0.114, 0.117 |
| 3 years | 0.069 | -0.105, 0.243 | -0.007 | -0.128, 0.113 |
| 5 years | 0.018 | -0.196, 0.232 | -0.026 | -0.175, 0.123 |
| 7 years | -0.032 | -0.310, 0.246 | -0.045 | -0.237, 0.147 |
| 8 years | -0.057 | -0.372, 0.257 | -0.054 | -0.270, 0.162 |
| **Male** |  |  |  |  |
| Average | **0.334** | **0.081**, **0.588** | 0.103 | -0.075, 0.282 |
| By age |  |  |  |  |
| 1 month | **0.332** | **0.081**, **0.583** | 0.102 | -0.074, 0.279 |
| 3 months | **0.327** | **0.081**, **0.573** | 0.101 | -0.073, 0.275 |
| 6 months | **0.320** | **0.080**, **0.560** | 0.099 | -0.072, 0.269 |
| 8 months | **0.313** | **0.078**, **0.547** | 0.096 | -0.071, 0.264 |
| 12 months | **0.305** | **0.075**, **0.536** | 0.094 | -0.071, 0.259 |
| 18 months | **0.291** | **0.066**, **0.515** | 0.089 | -0.073, 0.251 |
| 2 years | **0.276** | **0.053**, **0.499** | 0.084 | -0.078, 0.246 |
| 3 years | **0.247** | **0.015**, **0.478** | 0.075 | -0.094, 0.243 |
| 5 years | 0.188 | -0.101, 0.478 | 0.056 | -0.153, 0.264 |
| 7 years | 0.130 | -0.249, 0.509 | 0.037 | -0.232, 0.305 |
| 8 years | 0.101 | -0.329, 0.531 | 0.027 | -0.275, 0.329 |
| **Female** |  |  |  |  |
| Average | 0.008 | -0.269, 0.285 | -0.044 | -0.215, 0.126 |
| By age |  |  |  |  |
| 1 month | 0.005 | -0.269, 0.280 | -0.045 | -0.215, 0.124 |
| 3 months | 0.000 | -0.270, 0.270 | -0.048 | -0.214, 0.119 |
| 6 months | -0.008 | -0.272, 0.256 | -0.051 | -0.214, 0.113 |
| 8 months | -0.016 | -0.275, 0.243 | -0.054 | -0.215, 0.107 |
| 12 months | -0.024 | -0.278, 0.231 | -0.057 | -0.216, 0.102 |
| 18 months | -0.040 | -0.288, 0.209 | -0.063 | -0.220, 0.094 |
| 2 years | -0.055 | -0.303, 0.192 | -0.069 | -0.227, 0.089 |
| 3 years | -0.087 | -0.343, 0.169 | -0.082 | -0.249, 0.086 |
| 5 years | -0.151 | -0.466, 0.165 | -0.107 | -0.317, 0.104 |
| 7 years | -0.214 | -0.622, 0.194 | -0.132 | -0.404, 0.141 |
| 8 years | -0.246 | -0.707, 0.215 | -0.144 | -0.451, 0.163 |

All models are weighted for sociodemographic-lifestyle-reproductive factors (i.e., maternal age, parity, marital status, obstetric comorbidity index adapted from Bateman et al., maternal education, maternal gross yearly income, smoking and alcohol use in pregnancy, BMI at conception, history of abortions/miscarriages, folic acid intake, paternal age, paternal education, paternal social benefits, and paternal BMI), maternal psychiatric correlates (i.e., Lifetime History of Major Depression measured in Q1, SCL-5 measured in Q1 and Q3), comedication in pregnancy (i.e., opioid analgesics – ATC code N02A, benzodiazepines/z-hypnotics - ATC codes N05B and N05C, antipsychotics and mood stabilizers - ATC code N05A, and antiepileptics - ATC code N03A)

**Table S5**. Weighted differences in children’s BMI (in kg/m2) up to 8 years of age between prenatal antidepressant exposure groups (n=5,780 mother-child pairs, restricted to first pregnancy), overall and by child sex

|  | Antidepressant exposure | | | |
| --- | --- | --- | --- | --- |
|  | Continuers vs discontinuers | | Continuers vs unexposed | |
|  | beta | 95%CI | beta | 95%CI |
| **All children** |  |  |  |  |
| Average | 0.050 | -0.124, 0.223 | -0.009 | -0.126, 0.108 |
| By age |  |  |  |  |
| 1 month | 0.048 | -0.124, 0.220 | -0.010 | -0.126, 0.106 |
| 3 months | 0.045 | -0.124, 0.214 | -0.012 | -0.127, 0.103 |
| 6 months | 0.041 | -0.124, 0.206 | -0.015 | -0.127, 0.098 |
| 8 months | 0.037 | -0.125, 0.199 | -0.018 | -0.129, 0.093 |
| 12 months | 0.032 | -0.127, 0.192 | -0.021 | -0.130, 0.089 |
| 18 months | 0.024 | -0.133, 0.181 | -0.027 | -0.135, 0.081 |
| 2 years | 0.015 | -0.141, 0.172 | -0.033 | -0.141, 0.076 |
| 3 years | -0.002 | -0.165, 0.161 | -0.044 | -0.159, 0.070 |
| 5 years | -0.036 | -0.239, 0.167 | -0.068 | -0.211, 0.075 |
| 7 years | -0.070 | -0.334, 0.193 | -0.091 | -0.275, 0.092 |
| 8 years | -0.088 | -0.385, 0.210 | -0.103 | -0.310, 0.103 |
| **Male** |  |  |  |  |
| Average | 0.189 | -0.044, 0.422 | 0.015 | -0.149, 0.179 |
| By age |  |  |  |  |
| 1 month | 0.188 | -0.043, 0.418 | 0.014 | -0.149, 0.177 |
| 3 months | 0.186 | -0.041, 0.412 | 0.013 | -0.147, 0.174 |
| 6 months | 0.182 | -0.039, 0.403 | 0.011 | -0.146, 0.169 |
| 8 months | 0.178 | -0.038, 0.395 | 0.010 | -0.145, 0.164 |
| 12 months | 0.175 | -0.038, 0.388 | 0.008 | -0.145, 0.160 |
| 18 months | 0.168 | -0.040, 0.376 | 0.004 | -0.146, 0.154 |
| 2 years | 0.161 | -0.045, 0.367 | 0.001 | -0.150, 0.151 |
| 3 years | 0.147 | -0.068, 0.361 | -0.007 | -0.164, 0.151 |
| 5 years | 0.119 | -0.148, 0.385 | -0.021 | -0.216, 0.174 |
| 7 years | 0.090 | -0.257, 0.438 | -0.035 | -0.286, 0.215 |
| 8 years | 0.076 | -0.318, 0.470 | -0.043 | -0.325, 0.240 |
| **Female** |  |  |  |  |
| Average | -0.064 | -0.317, 0.190 | -0.031 | -0.190, 0.128 |
| By age |  |  |  |  |
| 1 month | -0.066 | -0.318, 0.186 | -0.033 | -0.191, 0.125 |
| 3 months | -0.071 | -0.319, 0.177 | -0.036 | -0.192, 0.120 |
| 6 months | -0.078 | -0.322, 0.166 | -0.040 | -0.193, 0.114 |
| 8 months | -0.086 | -0.326, 0.155 | -0.044 | -0.196, 0.108 |
| 12 months | -0.093 | -0.331, 0.145 | -0.048 | -0.198, 0.102 |
| 18 months | -0.108 | -0.343, 0.128 | -0.056 | -0.206, 0.094 |
| 2 years | -0.123 | -0.359, 0.114 | -0.065 | -0.217, 0.088 |
| 3 years | -0.152 | -0.403, 0.099 | -0.081 | -0.245, 0.082 |
| 5 years | -0.211 | -0.524, 0.102 | -0.114 | -0.322, 0.094 |
| 7 years | -0.270 | -0.673, 0.133 | -0.147 | -0.416, 0.122 |
| 8 years | -0.299 | -0.753, 0.154 | -0.164 | -0.466, 0.139 |

All models are weighted for sociodemographic-lifestyle-reproductive factors (i.e., maternal age, parity, marital status, obstetric comorbidity index adapted from Bateman et al., maternal education, maternal gross yearly income, smoking and alcohol use before pregnancy, BMI at conception, history of abortions/miscarriages, folic acid intake, paternal age, paternal education, paternal social benefits, and paternal BMI), maternal psychiatric correlates (i.e., Lifetime History of Major Depression measured in Q1), comedication before pregnancy (i.e., opioid analgesics – ATC code N02A, benzodiazepines/z-hypnotics - ATC codes N05B and N05C, antipsychotics and mood stabilizers - ATC code N05A, and antiepileptics - ATC code N03A) using inverse probability of treatment weighting.

**Table S6**. Weighted differences in children’s BMI (in kg/m2) up to 8 years of age between prenatal antidepressant exposure groups (n=5,768 mother-child pairs restricted to term born children), overall and by child sex

|  | Antidepressant exposure | | | |
| --- | --- | --- | --- | --- |
|  | Continuers vs discontinuers | | Continuers vs unexposed | |
|  | beta | 95%CI | beta | 95%CI |
| **All children** |  |  |  |  |
| Average | 0.050 | -0.123, 0.223 | -0.025 | -0.142, 0.092 |
| By age |  |  |  |  |
| 1 month | 0.048 | -0.123, 0.220 | -0.026 | -0.142, 0.090 |
| 3 months | 0.044 | -0.124, 0.213 | -0.028 | -0.142, 0.087 |
| 6 months | 0.039 | -0.126, 0.204 | -0.031 | -0.143, 0.081 |
| 8 months | 0.033 | -0.129, 0.194 | -0.034 | -0.144, 0.076 |
| 12 months | 0.027 | -0.132, 0.186 | -0.037 | -0.146, 0.071 |
| 18 months | 0.016 | -0.140, 0.171 | -0.044 | -0.151, 0.063 |
| 2 years | 0.004 | -0.151, 0.158 | -0.050 | -0.158, 0.057 |
| 3 years | -0.019 | -0.180, 0.141 | -0.063 | -0.176, 0.050 |
| 5 years | -0.066 | -0.265, 0.134 | -0.089 | -0.230, 0.053 |
| 7 years | -0.112 | -0.372, 0.148 | -0.114 | -0.297, 0.068 |
| 8 years | -0.135 | -0.429, 0.158 | -0.127 | -0.333, 0.079 |
| **Male** |  |  |  |  |
| Average | 0.204 | -0.032, 0.440 | 0.014 | -0.153, 0.181 |
| By age |  |  |  |  |
| 1 month | 0.202 | -0.032, 0.436 | 0.013 | -0.152, 0.178 |
| 3 months | 0.199 | -0.031, 0.429 | 0.011 | -0.151, 0.174 |
| 6 months | 0.194 | -0.030, 0.418 | 0.008 | -0.151, 0.168 |
| 8 months | 0.189 | -0.030, 0.408 | 0.006 | -0.151, 0.162 |
| 12 months | 0.184 | -0.031, 0.399 | 0.003 | -0.151, 0.157 |
| 18 months | 0.174 | -0.036, 0.383 | -0.003 | -0.154, 0.148 |
| 2 years | 0.164 | -0.044, 0.371 | -0.008 | -0.159, 0.142 |
| 3 years | 0.143 | -0.071, 0.358 | -0.020 | -0.177, 0.138 |
| 5 years | 0.103 | -0.164, 0.370 | -0.042 | -0.237, 0.153 |
| 7 years | 0.063 | -0.287, 0.412 | -0.065 | -0.317, 0.188 |
| 8 years | 0.042 | -0.354, 0.439 | -0.076 | -0.360, 0.209 |
| **Female** |  |  |  |  |
| Average | -0.074 | -0.320, 0.172 | -0.046 | -0.204, 0.113 |
| By age |  |  |  |  |
| 1 month | -0.077 | -0.321, 0.167 | -0.047 | -0.204, 0.110 |
| 3 months | -0.083 | -0.323, 0.157 | -0.050 | -0.205, 0.105 |
| 6 months | -0.092 | -0.328, 0.143 | -0.054 | -0.206, 0.098 |
| 8 months | -0.101 | -0.333, 0.130 | -0.058 | -0.208, 0.091 |
| 12 months | -0.111 | -0.339, 0.118 | -0.062 | -0.210, 0.086 |
| 18 months | -0.129 | -0.354, 0.097 | -0.071 | -0.217, 0.076 |
| 2 years | -0.147 | -0.373, 0.079 | -0.079 | -0.227, 0.069 |
| 3 years | -0.184 | -0.423, 0.055 | -0.096 | -0.254, 0.062 |
| 5 years | -0.257 | -0.556, 0.041 | -0.129 | -0.330, 0.071 |
| 7 years | -0.331 | -0.717, 0.056 | -0.163 | -0.424, 0.098 |
| 8 years | -0.367 | -0.803, 0.068 | -0.180 | -0.474, 0.115 |

All models are weighted for sociodemographic-lifestyle-reproductive factors (i.e., maternal age, parity, marital status, obstetric comorbidity index adapted from Bateman et al., maternal education, maternal gross yearly income, smoking and alcohol use in pregnancy, BMI at conception, history of abortions/miscarriages, folic acid intake, paternal age, paternal education, paternal social benefits, and paternal BMI), maternal psychiatric correlates (i.e., Lifetime History of Major Depression measured in Q1, SCL-5 measured in Q1 and Q3), comedication in pregnancy (i.e., opioid analgesics – ATC code N02A, benzodiazepines/z-hypnotics - ATC codes N05B and N05C, antipsychotics and mood stabilizers - ATC code N05A, and antiepileptics - ATC code N03A).

**Table S7**. Weighted differences in children’s BMI (in kg/m2) up to 8 years of age between prenatal antidepressant exposure groups (n=5,918 mother-child pairs in non-low-birth -weight children), overall and by child sex

|  | Antidepressant exposure | | | |
| --- | --- | --- | --- | --- |
|  | Continuers vs discontinuers | | Continuers vs unexposed | |
|  | beta | 95%CI | beta | 95%CI |
| **All children** |  |  |  |  |
| Average | 0.037 | -0.133, 0.207 | -0.029 | -0.143, 0.085 |
| By age |  |  |  |  |
| 1 month | 0.036 | -0.133, 0.204 | -0.030 | -0.143, 0.084 |
| 3 months | 0.034 | -0.132, 0.200 | -0.030 | -0.142, 0.082 |
| 6 months | 0.032 | -0.130, 0.194 | -0.031 | -0.141, 0.078 |
| 8 months | 0.029 | -0.129, 0.188 | -0.032 | -0.140, 0.076 |
| 12 months | 0.027 | -0.129, 0.183 | -0.033 | -0.139, 0.073 |
| 18 months | 0.022 | -0.131, 0.174 | -0.035 | -0.140, 0.070 |
| 2 years | 0.017 | -0.135, 0.169 | -0.037 | -0.142, 0.068 |
| 3 years | 0.007 | -0.151, 0.165 | -0.041 | -0.152, 0.070 |
| 5 years | -0.013 | -0.210, 0.185 | -0.048 | -0.188, 0.091 |
| 7 years | -0.033 | -0.290, 0.225 | -0.056 | -0.236, 0.125 |
| 8 years | -0.043 | -0.334, 0.249 | -0.060 | -0.263, 0.144 |
| **Male** |  |  |  |  |
| Average | 0.190 | -0.041, 0.420 | 0.019 | -0.141, 0.179 |
| By age |  |  |  |  |
| 1 month | 0.189 | -0.039, 0.417 | 0.019 | -0.140, 0.178 |
| 3 months | 0.188 | -0.036, 0.412 | 0.019 | -0.137, 0.176 |
| 6 months | 0.187 | -0.032, 0.405 | 0.020 | -0.133, 0.173 |
| 8 months | 0.185 | -0.028, 0.398 | 0.020 | -0.130, 0.170 |
| 12 months | 0.184 | -0.026, 0.393 | 0.020 | -0.128, 0.168 |
| 18 months | 0.181 | -0.023, 0.384 | 0.021 | -0.124, 0.166 |
| 2 years | 0.178 | -0.024, 0.379 | 0.022 | -0.123, 0.167 |
| 3 years | 0.172 | -0.037, 0.380 | 0.023 | -0.129, 0.175 |
| 5 years | 0.160 | -0.102, 0.421 | 0.026 | -0.165, 0.216 |
| 7 years | 0.148 | -0.197, 0.492 | 0.028 | -0.220, 0.276 |
| 8 years | 0.142 | -0.249, 0.532 | 0.030 | -0.250, 0.310 |
| **Female** |  |  |  |  |
| Average | -0.089 | -.0331, 0.154 | -0.065 | -0.221, 0.091 |
| By age |  |  |  |  |
| 1 month | -0.091 | -0.331, 0.150 | -0.066 | -0.221, 0.089 |
| 3 months | -0.095 | -0.332, 0.142 | -0.068 | -0.221, 0.085 |
| 6 months | -0.101 | -0.333, 0.132 | -0.071 | -0.221, 0.079 |
| 8 months | -0.107 | -0.336, 0.122 | -0.074 | -0.222, 0.074 |
| 12 months | -0.113 | -0.339, 0.113 | -0.077 | -0.223, 0.070 |
| 18 months | -0.125 | -0.348, 0.098 | -0.082 | -0.228, 0.063 |
| 2 years | -0.137 | -0.361, 0.086 | -0.088 | -0.235, 0.059 |
| 3 years | -0.162 | -0.398, 0.075 | -0.100 | -0.257, 0.058 |
| 5 years | -0.210 | -0.509, 0.088 | -0.122 | -0.323, 0.079 |
| 7 years | -0.259 | -0.646, 0.128 | -0.145 | -0.406, 0.116 |
| 8 years | -0.283 | -0.720, 0.154 | -0.156 | -0.450, 0.138 |

All models are weighted for sociodemographic-lifestyle-reproductive factors (i.e., maternal age, parity, marital status, obstetric comorbidity index adapted from Bateman et al., maternal education, maternal gross yearly income, smoking and alcohol use in pregnancy, BMI at conception, history of abortions/miscarriages, folic acid intake, paternal age, paternal education, paternal social benefits, and paternal BMI), maternal psychiatric correlates (i.e., Lifetime History of Major Depression measured in Q1, SCL-5 measured in Q1 and Q3), comedication in pregnancy (i.e., opioid analgesics – ATC code N02A, benzodiazepines/z-hypnotics - ATC codes N05B and N05C, antipsychotics and mood stabilizers - ATC code N05A, and antiepileptics - ATC code N03A)

**Table S8**. Weighted differences in children’s BMI (in kg/m2) up to 8 years of age between prenatal antidepressant exposure groups (n=6,084 mother-child pairs), overall and by child sex with adjustment for disease severity measured by the 5 items in the Hopkins Symptoms Checklist

|  | Antidepressant exposure | | | |
| --- | --- | --- | --- | --- |
|  | Continuers vs discontinuers | | Continuers vs unexposed | |
|  | beta | 95%CI | beta | 95%CI |
| **All children** |  |  |  |  |
| Average | 0.034 | -0.136, 0.205 | -0.027 | -0.143, 0.088 |
| By age |  |  |  |  |
| 1 month | 0.033 | -0.136, 0.202 | -0.028 | -0.143, 0.087 |
| 3 months | 0.030 | -0.136, 0.196 | -0.029 | -0.142, 0.084 |
| 6 months | 0.026 | -0.137, 0.188 | -0.031 | -0.142, 0.080 |
| 8 months | 0.021 | -0.138, 0.180 | -0.033 | -0.142, 0.076 |
| 12 months | 0.017 | -0.139, 0.173 | -0.035 | -0.143, 0.073 |
| 18 months | 0.008 | -0.145, 0.161 | -0.038 | -0.145, 0.068 |
| 2 years | -0.001 | -0.153, 0.151 | -0.042 | -0.149, 0.065 |
| 3 years | -0.018 | -0.177, 0.140 | -0.049 | -0.162, 0.063 |
| 5 years | -0.054 | -0.250, 0.143 | -0.064 | -0.203, 0.076 |
| 7 years | -0.089 | -0.344, 0.167 | -0.078 | -0.258, 0.101 |
| 8 years | -0.106 | -0.395, 0.182 | -0.086 | -0.287, 0.116 |
| **Male** |  |  |  |  |
| Average | 0.188 | -0.041, 0.416 | 0.018 | -0.143, 0.179 |
| By age |  |  |  |  |
| 1 month | 0.186 | -0.040, 0.413 | 0.018 | -0.142, 0.177 |
| 3 months | 0.184 | -0.039, 0.407 | 0.017 | -0.140, 0.174 |
| 6 months | 0.182 | -0.036, 0.400 | 0.015 | -0.139, 0.169 |
| 8 months | 0.180 | -0.033, 0.393 | 0.014 | -0.137, 0.165 |
| 12 months | 0.178 | -0.030, 0.386 | 0.013 | -0.136, 0.162 |
| 18 months | 0.174 | -0.029, 0.377 | 0.010 | -0.137, 0.157 |
| 2 years | 0.170 | -0.031, 0.371 | 0.007 | -0.139, 0.154 |
| 3 years | 0.162 | -0.046, 0.371 | 0.002 | -0.151, 0.155 |
| 5 years | 0.146 | -0.115, 0.408 | -0.009 | -0.198, 0.180 |
| 7 years | 0.131 | -0.213, 0.474 | -0.019 | -0.263, 0.224 |
| 8 years | 0.123 | -0.268, 0.513 | -0.025 | -0.299, 0.250 |
| **Female** |  |  |  |  |
| Average | -0.098 | -0.342, 0.146 | -0.060 | -0.218, 0.098 |
| By age |  |  |  |  |
| 1 month | -0.101 | -0.343, 0.142 | -0.060 | -0.217, 0.096 |
| 3 months | -0.106 | -0.344, 0.133 | -0.062 | -0.217, 0.092 |
| 6 months | -0.113 | -0.347, 0.121 | -0.065 | -0.217, 0.087 |
| 8 months | -0.121 | -0.351, 0.110 | -0.068 | -0.218, 0.082 |
| 12 months | -0.128 | -0.355, 0.099 | -0.070 | -0.219, 0.078 |
| 18 months | -0.143 | -0.367, 0.081 | -0.076 | -0.223, 0.071 |
| 2 years | -0.158 | -0.383, 0.066 | -0.081 | -0.230, 0.067 |
| 3 years | -0.189 | -0.426, 0.048 | -0.092 | -0.251, 0.067 |
| 5 years | -0.249 | -0.546, 0.048 | -0.114 | -0.315, 0.087 |
| 7 years | -0.310 | -0.694, 0.075 | -0.136 | -0.396, 0.124 |
| 8 years | -0.340 | -0.773, 0.094 | -0.147 | -0.440, 0.146 |

All models are weighted for sociodemographic-lifestyle-reproductive factors (i.e., maternal age, parity, marital status, obstetric comorbidity index adapted from Bateman et al., maternal education, maternal gross yearly income, smoking and alcohol use in pregnancy, BMI at conception, history of abortions/miscarriages, folic acid intake, paternal age, paternal education, paternal social benefits, and paternal BMI), maternal psychiatric correlates (i.e., Lifetime History of Major Depression measured in Q1, SCL-5 measured in Q1 and Q3), comedication in pregnancy (i.e., opioid analgesics – ATC code N02A, benzodiazepines/z-hypnotics - ATC codes N05B and N05C, antipsychotics and mood stabilizers - ATC code N05A, and antiepileptics - ATC code N03A)

**Table S9**. Weighted differences in children’s BMI (in kg/m2) up to 8 years of age between prenatal antidepressant exposure groups (n=1,913 mother-child pairs in having genetic data), overall and by child sex with additional adjustment for polygenic risk scores (continuous)

|  | Antidepressant exposure | | | |
| --- | --- | --- | --- | --- |
|  | Continuers vs discontinuers | | Continuers vs unexposed | |
|  | beta | 95%CI | beta | 95%CI |
| **All children** |  |  |  |  |
| Average | 0.102 | -0.218, 0.422 | -0.108 | -0.306, 0.090 |
| By age |  |  |  |  |
| 1 month | 0.102 | -0.215, 0.419 | -0.109 | -0.305, 0.088 |
| 3 months | 0.101 | -0.210, 0.412 | -0.109 | -0.302, 0.084 |
| 6 months | 0.100 | -0.203, 0.402 | -0.110 | -0.300, 0.079 |
| 8 months | 0.098 | -0.197, 0.394 | -0.112 | -0.298, 0.074 |
| 12 months | 0.097 | -0.192, 0.386 | -0.013 | -0.296, 0.071 |
| 18 months | 0.095 | -0.185, 0.374 | -0.115 | -0.296, 0.066 |
| 2 years | 0.092 | -0.182, 0.367 | -0.117 | -0.299, 0.064 |
| 3 years | 0.088 | -0.193, 0.368 | -0.122 | -0.314, 0.070 |
| 5 years | 0.078 | -0.265, 0.421 | -0.131 | -0.374, 0.111 |
| 7 years | 0.069 | -0.380, 0.517 | -0.141 | -0.456, 0.174 |
| 8 years | 0.064 | -0.446, 0.573 | -0.145 | -0.501, 0.210 |
| **Male** |  |  |  |  |
| Average | 0.098 | -0.341, 0.536 | -0.202 | -0.473, 0.069 |
| By age |  |  |  |  |
| 1 month | 0.099 | -0.335, 0.532 | -0.201 | -0.469, 0.068 |
| 3 months | 0.101 | -0.323, 0.524 | -0.199 | -0.462, 0.064 |
| 6 months | 0.103 | -0.307, 0.513 | -0.196 | -0.452, 0.060 |
| 8 months | 0.106 | -0.291, 0.504 | -0.193 | -0.443, 0.057 |
| 12 months | 0.109 | -0.278, 0.495 | -0.190 | -0.435, 0.055 |
| 18 months | 0.114 | -0.254, 0.483 | -0.184 | -0.423, 0.054 |
| 2 years | 0.120 | -0.238, 0.477 | -0.178 | -0.416, 0.059 |
| 3 years | 0.131 | -0.228, 0.489 | -0.167 | -0.416, 0.083 |
| 5 years | 0.153 | -0.290, 0.594 | -0.144 | -0.466, 0.178 |
| 7 years | 0.174 | -0.417, 0.766 | -0.121 | -0.549, 0.308 |
| 8 years | 0.185 | -0.493, 0.863 | -0.109 | -0.598, 0.379 |
| **Female** |  |  |  |  |
| Average | 0.057 | -0.399, 0.551 | -0.007 | -0.288, 0.273 |
| By age |  |  |  |  |
| 1 month | 0.053 | -0.398, 0.504 | -0.009 | -0.288, 0.270 |
| 3 months | 0.048 | -0.397, 0.493 | -0.013 | -0.289, 0.263 |
| 6 months | 0.040 | -0.395, 0.476 | -0.019 | -0.292, 0.254 |
| 8 months | 0.032 | -0.395, 0.460 | -0.024 | -0.295, 0.246 |
| 12 months | 0.025 | -0.396, 0.446 | -0.030 | -0.299, 0.239 |
| 18 months | 0.009 | -0.403, 0.421 | -0.041 | -0.311, 0.228 |
| 2 years | -0.006 | -0.415, 0.403 | -0.053 | -0.327, 0.221 |
| 3 years | -0.037 | -0.459, 0.385 | -0.076 | -0.369, 0.218 |
| 5 years | -0.099 | -0.609, 0.411 | -0.121 | -0.489, 0.247 |
| 7 years | -0.161 | -0.810, 0.489 | -0.168 | -0.635, 0.302 |
| 8 years | -0.192 | -0.923, 0.539 | -0.189 | -0.714, 0.334 |

All models are weighted for sociodemographic-lifestyle-reproductive factors (i.e., maternal age, parity, marital status, obstetric comorbidity index adapted from Bateman et al., maternal education, maternal gross yearly income, smoking and alcohol use in pregnancy, BMI at conception, history of abortions/miscarriages, folic acid intake, paternal age, paternal education, paternal social benefits, and paternal BMI), maternal psychiatric correlates (i.e., Lifetime History of Major Depression measured in Q1, SCL-5 measured in Q1 and Q3), comedication in pregnancy (i.e., opioid analgesics – ATC code N02A, benzodiazepines/z-hypnotics - ATC codes N05B and N05C, antipsychotics and mood stabilizers - ATC code N05A, and antiepileptics - ATC code N03A) and adjusted for polygenic risk scores fitted for depression in mothers, parental BMI and antidepressant response in continuous form.

**Table S10**. Weighted differences in children’s BMI (in kg/m2) up to 8 years of age between prenatal antidepressant exposure groups (n=1,913 mother-child pairs in having genetic data), overall and by child sex with additional adjustment for polygenic risk scores (z-score)

|  | Antidepressant exposure | | | |
| --- | --- | --- | --- | --- |
|  | Continuers vs discontinuers | | Continuers vs unexposed | |
|  | beta | 95%CI | beta | 95%CI |
| **All children** |  |  |  |  |
| Average | 0.102 | -0.218, 0.422 | -0.108 | -0.306, 0.090 |
| By age |  |  |  |  |
| 1 month | 0.102 | -0.215, 0.419 | -0.109 | -0.305, 0.088 |
| 3 months | 0.101 | -0.210, 0.412 | -0.109 | -0.302, 0.084 |
| 6 months | 0.100 | -0.203, 0.402 | -0.110 | -0.300, 0.079 |
| 8 months | 0.098 | -0.197, 0.393 | -0.112 | -0.298, 0.074 |
| 12 months | 0.097 | -0.192, 0.386 | -0.113 | -0.296, 0.071 |
| 18 months | 0.095 | -0.185, 0.374 | -0.115 | -0.296, 0.066 |
| 2 years | 0.092 | -0.182, 0.367 | -0.117 | -0.299, 0.064 |
| 3 years | 0.088 | -0.193, 0.368 | -0.122 | -0.314, 0.070 |
| 5 years | 0.078 | -0.265, 0.421 | -0.131 | -0.374, 0.111 |
| 7 years | 0.069 | -0.380, 0.517 | -0.141 | -0.456, 0.174 |
| 8 years | 0.064 | -0.446, 0.573 | -0.145 | -0.501, 0.210 |
| **Male** |  |  |  |  |
| Average | 0.098 | -0.341, 0.536 | -0.202 | -0.473, 0.069 |
| By age |  |  |  |  |
| 1 month | 0.099 | -0.335, 0.532 | -0.201 | -0.469, 0.068 |
| 3 months | 0.101 | -0.323, 0.524 | -0.199 | -0.462, 0.064 |
| 6 months | 0.103 | -0.307, 0.513 | -0.196 | -0.452, 0.060 |
| 8 months | 0.106 | -0.291, 0.504 | -0.193 | -0.443, 0.057 |
| 12 months | 0.109 | -0.278, 0.495 | -0.190 | -0.435, 0.055 |
| 18 months | 0.114 | -0.254, 0.483 | -0.184 | -0.423, 0.054 |
| 2 years | 0.120 | -0.238, 0.477 | -0.178 | -0.416, 0.059 |
| 3 years | 0.131 | -0.228, 0.489 | -0.169 | -0.416, 0.083 |
| 5 years | 0.153 | -0.290, 0.595 | -0.144 | -0.466, 0.178 |
| 7 years | 0.174 | -0.417, 0.766 | -0.121 | -0.549, 0.308 |
| 8 years | 0.185 | -0.493, 0.863 | -0.109 | -0.598, 0.379 |
| **Female** |  |  |  |  |
| Average | 0.056 | -0.399, 0.511 | -0.007 | -0.288, 0.273 |
| By age |  |  |  |  |
| 1 month | 0.053 | -0.398, 0.505 | -0.009 | -.0288, 0.270 |
| 3 months | 0.048 | -0.397, 0.493 | -0.013 | -0.289, 0.263 |
| 6 months | 0.040 | -0.395, 0.476 | -0.019 | -0.292, 0.254 |
| 8 months | 0.032 | -0.395, 0.460 | -0.024 | -0.295, 0.246 |
| 12 months | 0.028 | -0.396, 0.446 | -0.030 | -0.299, 0.239 |
| 18 months | 0.009 | -0.403, 0.421 | -0.041 | -0.311, 0.228 |
| 2 years | -0.006 | -0.415, 0.403 | -0.053 | -0.327, 0.221 |
| 3 years | -0.037 | -0.459, 0.385 | -0.076 | -0.369, 0.218 |
| 5 years | -0.099 | -0.609, 0.411 | -0.121 | -0.489, 0.247 |
| 7 years | -0.161 | -0.811, 0.489 | -0.168 | -0.635, 0.302 |
| 8 years | -0.192 | -0.922, 0.539 | -0.190 | -0.714, 0.335 |

All models are weighted for sociodemographic-lifestyle-reproductive factors (i.e., maternal age, parity, marital status, obstetric comorbidity index adapted from Bateman et al., maternal education, maternal gross yearly income, smoking and alcohol use in pregnancy, BMI at conception, history of abortions/miscarriages, folic acid intake, paternal age, paternal education, paternal social benefits, and paternal BMI), maternal psychiatric correlates (i.e., Lifetime History of Major Depression measured in Q1, SCL-5 measured in Q1 and Q3), comedication in pregnancy (i.e., opioid analgesics – ATC code N02A, benzodiazepines/z-hypnotics - ATC codes N05B and N05C, antipsychotics and mood stabilizers - ATC code N05A, and antiepileptics - ATC code N03A) and adjusted for polygenic risk scores fitted for depression in mothers, parental BMI and antidepressant response in form of z-scores.

**Table S11**. Weighted differences in children’s BMI (in kg/m2) up to 8 years of age between prenatal antidepressant exposure groups (n=418 mother-child pairs of those in 1^st^ quartile of polygenic risk score for maternal depression), overall and by child sex

|  | Antidepressant exposure | | | |
| --- | --- | --- | --- | --- |
|  | Continuers vs discontinuers | | Continuers vs unexposed | |
|  | beta | 95%CI | beta | 95%CI |
| **All children** |  |  |  |  |
| Average | -0.295 | -0.940, 0.349 | -0.141 | -0.582, 0.300 |
| By age |  |  |  |  |
| 1 month | -0.294 | -0.932, 0.342 | -0.140 | -0.578, 0.297 |
| 3 months | -0.292 | -0.916, 0.332 | -0.139 | -0.569, 0.291 |
| 6 months | -0.288 | -0.895, 0.318 | -0.137 | -0.556, 0.283 |
| 8 months | -0.285 | -0.876, 0.307 | -0.134 | -0.545, 0.277 |
| 12 months | -0.281 | -0.861, 0.299 | -0.132 | -0.536, 0.272 |
| 18 months | -0.274 | -0.841, 0.293 | -0.127 | -0.523, 0.268 |
| 2 years | -0.267 | -0.835, 0.301 | -0.123 | -0.516, 0.271 |
| 3 years | -0.253 | -0.863, 0.357 | -0.113 | -0.524, 0.297 |
| 5 years | -0.224 | -1.041, 0.592 | -0.095 | -0.607, 0.419 |
| 7 years | -0.196 | -1.300, 0.907 | -0.076 | -0.744, 0.592 |
| 8 years | -0.182 | -1.443, 1.079 | -0.067 | -0.823, 0.690 |
| **Male** |  |  |  |  |
| Average | 0.256 | -0.649, 1.160 | -0.044 | -0.754, 0.667 |
| By age |  |  |  |  |
| 1 month | 0.256 | -0.636, 1.147 | -0.038 | -0.740, 0.663 |
| 3 months | 0.256 | -0.611, 1.122 | -0.027 | -0.711, 0.658 |
| 6 months | 0.255 | -0.577, 1.088 | -0.010 | -0.671, 0.651 |
| 8 months | 0.255 | -0.548, 1.059 | 0.007 | -0.634, 0.647 |
| 12 months | 0.255 | -0.523, 1.033 | 0.024 | -0.599, 0.646 |
| 18 months | 0.255 | -0.490, 1.000 | 0.057 | -0.540, 0.654 |
| 2 years | 0.255 | -0.481, 0.991 | 0.091 | -0.495, 0.677 |
| 3 years | 0.254 | -0.535, 1.044 | 0.158 | -0.452, 0.768 |
| 5 years | 0.254 | -0.854, 1.361 | 0.292 | -0.513, 1.100 |
| 7 years | 0.253 | -1.302, 1.807 | 0.427 | -0.676, 1.530 |
| 8 years | 0.252 | -1.545, 2.050 | 0.494 | -0.774, 1.763 |
| **Female** |  |  |  |  |
| Average | **-0.935** | **-1.844**, **-0.027** | -0.170 | -0.656, 0.315 |
| By age |  |  |  |  |
| 1 month | **-0.931** | **-1.834**, **-0.028** | -0.174 | -0.657, 0.309 |
| 3 months | **-0.922** | **-1.814**, **-0.030** | -0.180 | -0.660, 0.299 |
| 6 months | **-0.909** | **-1.787**, **-0.031** | -0.191 | -0.665, 0.284 |
| 8 months | **-0.896** | **-1.762**, **-0.030** | -0.201 | -0.671, 0.270 |
| 12 months | **-0.883** | **-1.740**, **-0.026** | -0.211 | -0.679, 0.257 |
| 18 months | **-0.857** | **-1.703**, **-0.011** | -0.231 | -0.699, 0.236 |
| 2 years | -0.831 | -1.677, 0.015 | -0.252 | -0.723, 0.220 |
| 3 years | -0.779 | -1.656, 0.099 | -0.292 | -0.787, 0.203 |
| 5 years | -0.674 | -1.722, 0.374 | -0.373 | -0.963, 0.215 |
| 7 years | -0.570 | -1.880, 0.741 | -0.455 | -1.180, 0.271 |
| 8 years | -0.517 | -1.980, 0.944 | -0.495 | -1.300, 0.308 |

All models are weighted for sociodemographic-lifestyle-reproductive factors (i.e., maternal age, parity, marital status, obstetric comorbidity index adapted from Bateman et al., maternal education, maternal gross yearly income, smoking and alcohol use in pregnancy, BMI at conception, history of abortions/miscarriages, folic acid intake, paternal age, paternal education, paternal social benefits, and paternal BMI), maternal psychiatric correlates (i.e., Lifetime History of Major Depression measured in Q1, SCL-5 measured in Q1 and Q3), comedication in pregnancy (i.e., opioid analgesics – ATC code N02A, benzodiazepines/z-hypnotics - ATC codes N05B and N05C, antipsychotics and mood stabilizers - ATC code N05A, and antiepileptics - ATC code N03A)

**Table S12**. Weighted differences in children’s BMI (in kg/m2) up to 8 years of age between prenatal antidepressant exposure groups (n=551 mother-child pairs in those in 4^th^ quartile of polygenic risk score for maternal depression), overall and by child sex

|  | Antidepressant exposure | | | |
| --- | --- | --- | --- | --- |
|  | Continuers vs discontinuers | | Continuers vs unexposed | |
|  | beta | 95%CI | beta | 95%CI |
| **All children** |  |  |  |  |
| Average | 0.108 | -0.459, 0.675 | 0.014 | -0.295, 0.323 |
| By age |  |  |  |  |
| 1 month | 0.116 | -0.446, 0.678 | 0.011 | -0.296, 0.317 |
| 3 months | 0.133 | -0.420, 0.685 | 0.004 | -0.299, 0.307 |
| 6 months | 0.157 | -0.383, 0.697 | -0.006 | -0.305, 0.292 |
| 8 months | 0.181 | -0.348, 0.711 | -0.017 | -0.312, 0.279 |
| 12 months | 0.206 | -0.315, 0.726 | -0.027 | -0.322, 0.268 |
| 18 months | 0.254 | -0.254, 0.763 | -0.048 | -0.347, 0.252 |
| 2 years | 0.303 | -0.202, 0.808 | -0.068 | -0.381, 0.244 |
| 3 years | 0.400 | -0.123, 0.924 | -0.109 | -0.466, 0.248 |
| 5 years | 0.595 | -0.050, 1.240 | -0.192 | -0.686, 0.303 |
| 7 years | 0.790 | -0.045, 1.624 | -0.274 | -0.936, 0.388 |
| 8 years | 0.887 | -0.056, 1.830 | -0.315 | -1.066, 0.436 |
| **Male** |  |  |  |  |
| Average | -0.570 | -1.250, 0.109 | -0.337 | -0.687, 0.012 |
| By age |  |  |  |  |
| 1 month | -0.555 | -1.229, 0.120 | -0.340 | -0.687, 0.007 |
| 3 months | -0.524 | -1.189, 0.141 | -0.345 | -0.687, -0.004 |
| 6 months | -0.478 | -1.130, 0.174 | -0.354 | -0.689, -0.018 |
| 8 months | -0.432 | -1.072, 0.209 | -0.362 | -0.693, -0.031 |
| 12 months | -0.386 | -1.016, 0.245 | -0.370 | -0.697, -0.043 |
| 18 months | -0.293 | -0.909, 0.322 | -0.387 | -0.712, -0.062 |
| 2 years | -0.201 | -0.809, 0.407 | -0.403 | -0.732, -0.075 |
| 3 years | -0.017 | -0.632, 0.598 | -0.436 | -0.789, -0.084 |
| 5 years | 0.352 | -0.357, 1.061 | -0.503 | -0.954, -0.051 |
| 7 years | 0.721 | -0.158, 1.600 | -0.569 | -1.157, 0.020 |
| 8 years | 0.905 | -0.075, 1.886 | -0.602 | -1.266, 0.062 |
| **Female** |  |  |  |  |
| Average | 0.738 | -0.054, 1.530 | 0.389 | -0.084, 0.861 |
| By age |  |  |  |  |
| 1 month | 0.740 | -0.048, 1.528 | 0.384 | -0.085, 0.854 |
| 3 months | 0.744 | -0.036, 1.524 | 0.375 | -0.090, 0.841 |
| 6 months | 0.749 | -0.022, 1.520 | 0.362 | -0.100, 0.824 |
| 8 months | 0.755 | -0.010, 1.520 | 0.348 | -0.113, 0.810 |
| 12 months | 0.760 | -0.002, 1.523 | 0.335 | -0.131, 0.800 |
| 18 months | 0.772 | 0.005, 1.538 | 0.308 | -0.176, 0.792 |
| 2 years | 0.783 | -0.000, 1.566 | 0.281 | -0.233, 0.795 |
| 3 years | 0.805 | -0.045, 1.655 | 0.227 | -0.378, 0.833 |
| 5 years | 0.849 | -0.236, 1.934 | 0.119 | -0.737, 0.796 |
| 7 years | 0.894 | -0.501, 2.288 | 0.012 | -1.136, 1.159 |
| 8 years | 0.916 | -0.648, 2.480 | -0.042 | -1.342, 1.258 |

All models are weighted for sociodemographic-lifestyle-reproductive factors (i.e., maternal age, parity, marital status, obstetric comorbidity index adapted from Bateman et al., maternal education, maternal gross yearly income, smoking and alcohol use in pregnancy, BMI at conception, history of abortions/miscarriages, folic acid intake, paternal age, paternal education, paternal social benefits, and paternal BMI), maternal psychiatric correlates (i.e., Lifetime History of Major Depression measured in Q1, SCL-5 measured in Q1 and Q3), comedication in pregnancy (i.e., opioid analgesics – ATC code N02A, benzodiazepines/z-hypnotics - ATC codes N05B and N05C, antipsychotics and mood stabilizers - ATC code N05A, and antiepileptics - ATC code N03A)

**Table S13**. Weighted differences in children’s BMI (in kg/m2) up to 8 years of age between prenatal antidepressant exposure groups (n=475 mother-child pairs in those in 1^st^ quartile of polygenic score for maternal BMI), overall and by child sex

|  | Antidepressant exposure | | | |
| --- | --- | --- | --- | --- |
|  | Continuers vs discontinuers | | Continuers vs unexposed | |
|  | beta | 95%CI | beta | 95%CI |
| **All children** |  |  |  |  |
| Average | -0.377 | -1.027, 0.274 | -0.255 | -0.592, 0.082 |
| By age |  |  |  |  |
| 1 month | -0.366 | -1.011, 0.280 | -0.250 | -0.586, 0.085 |
| 3 months | -0.345 | -0.980, 0.291 | -0.241 | -0.573, 0.091 |
| 6 months | -0.313 | -0.934, 0.309 | -0.227 | -0.554, 0.100 |
| 8 months | -0.281 | -0.889, 0.328 | -0.213 | -0.537, 0.110 |
| 12 months | -0.249 | -0.846, 0.348 | -0.199 | -0.520, 0.121 |
| 18 months | -0.185 | 0.764, 0.394 | -0.172 | -0.491, 0.148 |
| 2 years | -0.121 | -0.689, 0.446 | -0.144 | -0.467, 0.179 |
| 3 years | 0.007 | -0.558, 0.571 | -0.088 | -0.431, 0.255 |
| 5 years | 0.262 | -0.377, 0.901 | 0.023 | -0.402, 0.448 |
| 7 years | 0.517 | -0.274, 1.308 | 0.134 | -0.406, 0.674 |
| 8 years | 0.645 | -0.239, 1.529 | 0.190 | -0.414, 0.794 |
| **Male** |  |  |  |  |
| Average | **-0.816** | **-1.555**, **-0.076** | -0.368 | -0.814, 0.776 |
| By age |  |  |  |  |
| 1 month | **-0.798** | **-1.534**, **-0.063** | -0.365 | -0.809, 0.080 |
| 3 months | **-0.764** | **-1.492**, **-0.036** | -0.358 | -0.801, 0.085 |
| 6 months | -0.712 | -1.430, 0.006 | -0.348 | -0.790, 0.094 |
| 8 months | -0.660 | -1.370, 0.049 | -0.338 | -0.780, 0.105 |
| 12 months | -0.608 | -1.311, 0.094 | -0.327 | -0.773, 0.118 |
| 18 months | -0.505 | -1.200, 0.190 | -0.307 | -0.763, 0.149 |
| 2 years | -0.401 | -1.096, 0.293 | -0.287 | -0.760, 0.186 |
| 3 years | -0.194 | -0.910, 0.522 | -0.246 | -0.770, 0.277 |
| 5 years | 0.220 | -0.614, 1.054 | -0.165 | -0.838, 0.507 |
| 7 years | 0.635 | -0.388, 1.657 | -0.084 | -0.940, 0.772 |
| 8 years | 0.842 | -0.290, 1.974 | -0.043 | -0.997, 0.911 |
| **Female** |  |  |  |  |
| Average | 0.294 | -0.737, 1.324 | -0.058 | -0.535, 0.419 |
| By age |  |  |  |  |
| 1 month | 0.295 | -0.730, 1.319 | -0.054 | -0.528, 0.419 |
| 3 months | 0.296 | -0.717, 1.309 | -0.047 | -0.514, 0.420 |
| 6 months | 0.299 | -0.699, 1.296 | -0.035 | -0.495, 0.424 |
| 8 months | 0.301 | -0.683, 1.285 | -0.024 | -0.476, 0.429 |
| 12 months | 0.303 | -0.668, 1.275 | -0.012 | -0.460, 0.345 |
| 18 months | 0.308 | -0.644, 1.260 | 0.011 | -0.430, 0.452 |
| 2 years | 0.313 | -0.628, 1.253 | 0.033 | -0.407, 0.474 |
| 3 years | 0.322 | -0.617, 1.262 | 0.079 | -0.380, 0.538 |
| 5 years | 0.341 | -0.685, 1.368 | 0.171 | -0.385, 0.726 |
| 7 years | 0.360 | -0.847, 1.567 | 0.262 | -0.440, 0.964 |
| 8 years | 0.370 | -0.951, 1.691 | 0.308 | -0.478, 1.094 |

All models are weighted for sociodemographic-lifestyle-reproductive factors (i.e., maternal age, parity, marital status, obstetric comorbidity index adapted from Bateman et al., maternal education, maternal gross yearly income, smoking and alcohol use in pregnancy, BMI at conception, history of abortions/miscarriages, folic acid intake, paternal age, paternal education, paternal social benefits, and paternal BMI), maternal psychiatric correlates (i.e., Lifetime History of Major Depression measured in Q1, SCL-5 measured in Q1 and Q3), comedication in pregnancy (i.e., opioid analgesics – ATC code N02A, benzodiazepines/z-hypnotics - ATC codes N05B and N05C, antipsychotics and mood stabilizers - ATC code N05A, and antiepileptics - ATC code N03A)

**Table S14**. Weighted differences in children’s BMI (in kg/m2) up to 8 years of age between prenatal antidepressant exposure groups (n=504 mother-child pairs in those in 4^th^ quartile of polygenic score for maternal BMI), overall and by child sex

|  | Antidepressant exposure | | | |
| --- | --- | --- | --- | --- |
|  | Continuers vs discontinuers | | Continuers vs unexposed | |
|  | beta | 95%CI | beta | 95%CI |
| **All children** |  |  |  |  |
| Average | 0.389 | -0.179, 0.956 | 0.030 | -0.361, 0.420 |
| By age |  |  |  |  |
| 1 month | 0.382 | -0.177, 0.940 | 0.228 | -0.364, 0.410 |
| 3 months | 0.368 | -0.175, 0.911 | 0.009 | -0.371, 0.389 |
| 6 months | 0.348 | -0.175, 0.870 | -0.101 | -0.382, 0.360 |
| 8 months | 0.327 | -0.180, 0.834 | -0.031 | -0.396, 0.333 |
| 12 months | 0.307 | -0.190, 0.803 | -0.051 | -0.411, 0.308 |
| 18 months | 0.266 | -0.226, 0.757 | -0.092 | -0.449, 0.265 |
| 2 years | 0.225 | -0.284, 0.734 | -0.132 | -0.496, 0.231 |
| 3 years | 0.143 | -0.457, 0.743 | -0.213 | -0.614, 0.188 |
| 5 years | -0.020 | -0.929, 0.888 | -0.375 | -0.919, 0.169 |
| 7 years | -0.184 | -1.465, 1.097 | -0.537 | -1.268, 0.194 |
| 8 years | -0.266 | -1.743, 1.211 | -0.618 | -1.451, 0.214 |
| **Male** |  |  |  |  |
| Average | **0.810** | **0.091**, **1.529** | -0.012 | -0.445, 0.420 |
| By age |  |  |  |  |
| 1 month | **0.801** | **0.096**, **1.507** | -0.020 | -0.449, 0.410 |
| 3 months | **0.785** | **0.104**, **1.465** | -0.034 | -0.457, 0.389 |
| 6 months | **0.759** | **0.112**, **1.406** | -0.056 | -0.471, 0.359 |
| 8 months | **0.734** | **0.113**, **1.354** | -0.078 | -0.488, 0.333 |
| 12 months | **0.708** | **0.108**, **1.309** | -0.100 | -0.507, 0.308 |
| 18 months | **0.657** | **0.073**, **1.241** | -0.143 | -0.553, 0.267 |
| 2 years | **0.606** | **0.006**, **1.207** | -0.187 | -0.610, 0.236 |
| 3 years | 0.505 | -0.215, 1.225 | -0.274 | -0.749, 0.201 |
| 5 years | 0.301 | -0.841, 1.443 | -0.448 | -1.096, 0.200 |
| 7 years | 0.097 | -1.551, 1.746 | -0.622 | -1.149, 0.244 |
| 8 years | -0.004 | -1.917, 1.908 | -0.709 | -1.692, 0.273 |
| **Female** |  |  |  |  |
| Average | -0.010 | -0.898, 0.878 | 0.108 | -0.516, 0.733 |
| By age |  |  |  |  |
| 1 month | -0.019 | -0.900, 0.861 | 0.102 | -0.517, 0.720 |
| 3 months | -0.037 | -0.904, 0.830 | 0.089 | -0.518, 0.696 |
| 6 months | -0.065 | -0.917, 0.787 | 0.070 | -0.523, 0.662 |
| 8 months | -0.092 | -0.935, 0.750 | 0.050 | -0.532, 0.633 |
| 12 months | -0.120 | -0.960, 0.720 | 0.031 | -0.544, 0.606 |
| 18 months | -0.175 | -1.030, 0.680 | -0.007 | -0.580, 0.565 |
| 2 years | -0.230 | -1.124, 0.664 | -0.046 | -0.631, 0.539 |
| 3 years | -0.340 | -1.375, 0.694 | -0.128 | -0.775, 0.529 |
| 5 years | -0.560 | -2.023, 0.903 | -0.277 | -1.173, 0.619 |
| 7 years | -0.780 | -2.761, 1.200 | -0.430 | -1.640, 0.779 |
| 8 years | -0.891 | -3.143, 1.362 | -0.507 | -1.886, 0.871 |

All models are weighted for sociodemographic-lifestyle-reproductive factors (i.e., maternal age, parity, marital status, obstetric comorbidity index adapted from Bateman et al., maternal education, maternal gross yearly income, smoking and alcohol use in pregnancy, BMI at conception, history of abortions/miscarriages, folic acid intake, paternal age, paternal education, paternal social benefits, and paternal BMI), maternal psychiatric correlates (i.e., Lifetime History of Major Depression measured in Q1, SCL-5 measured in Q1 and Q3), comedication in pregnancy (i.e., opioid analgesics – ATC code N02A, benzodiazepines/z-hypnotics - ATC codes N05B and N05C, antipsychotics and mood stabilizers - ATC code N05A, and antiepileptics - ATC code N03A)

**Table S15**. Weighted differences in children’s BMI (in kg/m2) up to 8 years of age between prenatal antidepressant exposure groups (n=460 mother-child pairs in those in 1^st^ quartile of polygenic score for paternal BMI), overall and by child sex

|  | Antidepressant exposure | | | |
| --- | --- | --- | --- | --- |
|  | Continuers vs discontinuers | | Continuers vs unexposed | |
|  | beta | 95%CI | beta | 95%CI |
| **All children** |  |  |  |  |
| Average | -0.188 | -0.886, 0.510 | **-0.355** | **-0.703**, **-0.007** |
| By age |  |  |  |  |
| 1 month | -0.177 | -0.867, 0.513 | **-0.347** | **-0.693**, **-0.001** |
| 3 months | -0.155 | -0.829, 0.519 | -0.332 | -0.674, 0.009 |
| 6 months | -0.122 | -0.774, 0.530 | -0.310 | -0.646, 0.026 |
| 8 months | -0.089 | -0.721, 0.543 | -0.287 | -0.620, 0.045 |
| 12 months | -0.056 | -0.670, 0.557 | -0.265 | -0.595, 0.065 |
| 18 months | 0.010 | -0.572, 0.593 | -0.220 | -0.550, 0.110 |
| 2 years | 0.076 | -0.485, 0.638 | -0.175 | -0.512, 0.162 |
| 3 years | 0.209 | -0.344, 0.761 | -0.085 | -0.452, 0.282 |
| 5 years | 0.473 | -0.188, 1.134 | 0.095 | -0.383, 0.572 |
| 7 years | 0.738 | -0.141, 1.616 | 0.274 | -0.349, 0.898 |
| 8 years | 0.870 | -0.138, 1.878 | 0.364 | -0.339, 1.067 |
| **Male** |  |  |  |  |
| Average | -0.716 | -1.623, 0.192 | **-0.640** | **-1.076**, **-0.203** |
| By age |  |  |  |  |
| 1 month | -0.694 | -1.589, 0.201 | **-0.628** | **-1.063**, **-0.193** |
| 3 months | -0.651 | -1.522, 0.220 | **-0.605** | **-1.037**, **-0.173** |
| 6 months | -0.687 | -1.424, 0.251 | **-0.570** | **-1.000**, **-0.139** |
| 8 months | -0.522 | -1.330, 0.285 | **-0.534** | **-0.967**, **-0.102** |
| 12 months | -0.458 | -1.239, 0.323 | **-0.499** | **-0.937**, **-0.061** |
| 18 months | -0.329 | -1.069, 0.411 | -0.429 | -0.885, 0.027 |
| 2 years | -0.200 | -0.918, 0.517 | -0.359 | -0.844, 0.127 |
| 3 years | 0.058 | -0.678, 0.793 | -0.218 | -0.787, 0.351 |
| 5 years | 0.573 | -0.402, 1.549 | 0.063 | -0.731, 0.857 |
| 7 years | 1.089 | -0.268, 2.446 | 0.344 | -0.709, 1.398 |
| 8 years | 1.347 | -0.224, 2.918 | 0.485 | -0.705, 1.674 |
| **Female** |  |  |  |  |
| Average | 0.463 | -0.496, 1.422 | -0.035 | -0.544, 0.473 |
| By age |  |  |  |  |
| 1 month | 0.460 | -0.493, 1.414 | -0.032 | -0.537, 0.473 |
| 3 months | 0.455 | -0.487, 1.398 | -0.025 | -0.523, 0.472 |
| 6 months | 0.448 | -0.479, 1.375 | -0.016 | -0.503, 0.473 |
| 8 months | 0.440 | -0.473, 1.354 | -0.005 | -0.485, 0.475 |
| 12 months | 0.433 | -0.469, 1.335 | 0.005 | -0.469, 0.479 |
| 18 months | 0.418 | -0.468, 1.304 | 0.025 | -0.441, 0.491 |
| 2 years | 0.403 | -0.476, 1.282 | 0.045 | -0.421, 0.512 |
| 3 years | 0.373 | -0.521, 1.266 | 0.086 | -0.402, 0.574 |
| 5 years | 0.313 | -0.711, 1.337 | 0.167 | -0.436, 0.768 |
| 7 years | 0.253 | -0.998, 1.503 | 0.248 | -0.524, 1.019 |
| 8 years | 0.223 | -1.163, 1.609 | 0.288 | -0.580, 1.156 |

All models are weighted for sociodemographic-lifestyle-reproductive factors (i.e., maternal age, parity, marital status, obstetric comorbidity index adapted from Bateman et al., maternal education, maternal gross yearly income, smoking and alcohol use in pregnancy, BMI at conception, history of abortions/miscarriages, folic acid intake, paternal age, paternal education, paternal social benefits, and paternal BMI), maternal psychiatric correlates (i.e., Lifetime History of Major Depression measured in Q1, SCL-5 measured in Q1 and Q3), comedication in pregnancy (i.e., opioid analgesics – ATC code N02A, benzodiazepines/z-hypnotics - ATC codes N05B and N05C, antipsychotics and mood stabilizers - ATC code N05A, and antiepileptics - ATC code N03A)

**Table S16**. Weighted differences in children’s BMI (in kg/m2) up to 8 years of age between prenatal antidepressant exposure groups (n=499 mother-child pairs in those in 4th quartile of polygenic risk score for paternal BMI), overall and by child sex

|  | Antidepressant exposure | | | |
| --- | --- | --- | --- | --- |
|  | Continuers vs discontinuers | | Continuers vs unexposed | |
|  | beta | 95%CI | beta | 95%CI |
| **All children** |  |  |  |  |
| Average | 0.246 | -0.277, 0.769 | -0.058 | -0.450, 0.338 |
| By age |  |  |  |  |
| 1 month | 0.255 | -0.261, 0.771 | -0.056 | -0.444, 0.332 |
| 3 months | 0.273 | -0.229, 0.775 | -0.053 | -0.433, 0.327 |
| 6 months | 0.301 | -0.182, 0.784 | -0.047 | -0.417, 0.323 |
| 8 months | 0.328 | -0.139, 0.796 | -0.042 | -0.402, 0.319 |
| 12 months | 0.356 | -0.099, 0.811 | -0.036 | -0.389, 0.317 |
| 18 months | 0.411 | -0.029, 0.085 | -0.025 | -0.368, 0.318 |
| 2 years | 0.466 | 0.025, 0.907 | -0.014 | -0.354, 0.326 |
| 3 years | 0.576 | 0.091, 1.062 | 0.009 | -0.347, 0.365 |
| 5 years | 0.796 | 0.103, 1.489 | 0.053 | -0.405, 0.511 |
| 7 years | 1.017 | 0.047, 1.987 | 0.098 | -0.513, 0.709 |
| 8 years | 1.127 | 0.008, 2.246 | 0.120 | -0.576, 0.816 |
| **Male** |  |  |  |  |
| Average | 0.579 | -0.057, 1.215 | -0.079 | -0.491, 0.333 |
| By age |  |  |  |  |
| 1 month | 0.586 | -0.041, 1.212 | -0.076 | -0.486, 0.334 |
| 3 months | 0.598 | -0.010, 1.206 | -0.070 | -0.477, 0.336 |
| 6 months | 0.617 | 0.333, 1.200 | -0.062 | -0.464, 0.340 |
| 8 months | 0.635 | 0.072, 1.199 | -0.054 | -0.453, 0.345 |
| 12 months | 0.654 | 0.106, 1.202 | -0.046 | -0.443, 0.352 |
| 18 months | 0.692 | 0.158, 1.226 | -0.029 | -0.429, 0.370 |
| 2 years | 0.729 | 0.186, 1.272 | -0.013 | -0.420, 0.395 |
| 3 years | 0.804 | 0.182, 1.427 | 0.020 | -0.420, 0.460 |
| 5 years | 0.954 | 0.022, 1.886 | 0.086 | -0.467, 0.639 |
| 7 years | 1.104 | -0.217, 2.425 | 0.152 | -0.553, 0.856 |
| 8 years | 1.179 | -0.347, 2.706 | 0.185 | -0.603, 0.972 |
| **Female** |  |  |  |  |
| Average | -0.102 | -0.953, 0.750 | -0.017 | -0.670, 0.637 |
| By age |  |  |  |  |
| 1 month | -0.090 | -0.930, 0.750 | -0.016 | -0.661, 0.629 |
| 3 months | -0.067 | -0.885, 0.752 | -0.015 | -0.644, 0.614 |
| 6 months | -0.032 | -0.821, 0.758 | -0.013 | -0.619, 0.594 |
| 8 months | 0.003 | -0.761, 0.768 | -0.011 | -0.597, 0.576 |
| 12 months | 0.038 | -0.706, 0.782 | -0.009 | -0.578, 0.561 |
| 18 months | 0.108 | -0.610, 0.826 | -0.005 | -0.549, 0.539 |
| 2 years | 0.178 | -0.535, 0.891 | -0.001 | -0.533, 0.531 |
| 3 years | 0.318 | -0.449, 1.084 | 0.007 | -0.544, 0.559 |
| 5 years | 0.597 | -0.462, 1.657 | 0.023 | -0.706, 0.753 |
| 7 years | 0.877 | -0.592, 2.346 | 0.040 | -0.964, 1.043 |
| 8 years | 1.016 | -0.676, 2.708 | 0.048 | -1.109, 1.204 |

All models are weighted for sociodemographic-lifestyle-reproductive factors (i.e., maternal age, parity, marital status, obstetric comorbidity index adapted from Bateman et al., maternal education, maternal gross yearly income, smoking and alcohol use in pregnancy, BMI at conception, history of abortions/miscarriages, folic acid intake, paternal age, paternal education, paternal social benefits, and paternal BMI), maternal psychiatric correlates (i.e., Lifetime History of Major Depression measured in Q1, SCL-5 measured in Q1 and Q3), comedication in pregnancy (i.e., opioid analgesics – ATC code N02A, benzodiazepines/z-hypnotics - ATC codes N05B and N05C, antipsychotics and mood stabilizers - ATC code N05A, and antiepileptics - ATC code N03A)

**Table S17**. Weighted differences in children’s BMI (in kg/m2) up to 8 years of age between prenatal antidepressant exposure groups (n=475 mother-child pairs in those in 1^st^ quartile of polygenic score for maternal antidepressant response), overall and by child sex

|  | Antidepressant exposure | | | |
| --- | --- | --- | --- | --- |
|  | Continuers vs discontinuers | | Continuers vs unexposed | |
|  | beta | 95%CI | beta | 95%CI |
| **All children** |  |  |  |  |
| Average | -0.273 | -0.851, 0.305 | -0.314 | -0.748, 0.120 |
| By age |  |  |  |  |
| 1 month | -0.277 | -0.851, 0.297 | -0.315 | -0.756, 0.117 |
| 3 months | -0.285 | -0.853, 0.282 | -0.315 | -0.742, 0.111 |
| 6 months | -0.297 | -0.856, 0.262 | -0.317 | -0.736, 0.103 |
| 8 months | -0.310 | -0.862, 0.243 | -0.318 | -0.733, 0.097 |
| 12 months | -0.322 | -0.870, 0.226 | -0.319 | -0.730, 0.093 |
| 18 months | -0.346 | -0.892, 0.200 | -0.321 | -0.730, 0.088 |
| 2 years | -0.370 | -0.922, 0.181 | -0.324 | -0.736, 0.089 |
| 3 years | -0.419 | -1.007, 0.169 | -0.328 | -0.763, 0.107 |
| 5 years | -0.516 | -1.252, 0.219 | -0.338 | -0.873, 0.197 |
| 7 years | -0.614 | -1.555, 0.328 | -0.347 | -1.025, 0.331 |
| 8 years | -0.662 | -1.718, 0.393 | -0.352 | -1.111, 0.406 |
| **Male** |  |  |  |  |
| Average | -0.087 | -0.742, 0.568 | -0.372 | -1.027, 0.276 |
| By age |  |  |  |  |
| 1 month | -0.093 | -0.745, 0.559 | -0.373 | -1.021, 0.276 |
| 3 months | -0.106 | -0.753, 0.541 | -0.373 | -1.011, 0.266 |
| 6 months | -0.124 | -0.767, 0.518 | -0.373 | -0.997, 0.250 |
| 8 months | -0.143 | -0.783, 0.497 | -0.374 | -0.983, 0.236 |
| 12 months | -0.162 | -0.803, 0.479 | -0.374 | -0.972, 0.224 |
| 18 months | -0.199 | -0.849, 0.450 | -0.375 | -0.953, 0.204 |
| 2 years | -0.237 | -0.905 0.432 | -0.376 | -0.942, 0.191 |
| 3 years | -0.312 | -1.005, 0.422 | -0.377 | -0.941, 0.187 |
| 5 years | -0.461 | -1.403, 0.480 | -0.380 | -1.026, 0.265 |
| 7 years | -0.611 | -1.816, 0.595 | -0.383 | -1.191, 0.424 |
| 8 years | -0.686 | -2.034, 0.663 | -0.385 | -1.291, 0.522 |
| **Female** |  |  |  |  |
| Average | -0.389 | -1.354, 0.577 | -0.115 | -0.681, 0.456 |
| By age |  |  |  |  |
| 1 month | -0.392 | -1.350, 0.297 | -0.015 | -0.681, 0.452 |
| 3 months | -0.398 | -1.341, 0.282 | -0.120 | -0.682, 0.443 |
| 6 months | -0.407 | -1.332, 0.262 | -0.127 | -0.685, 0.431 |
| 8 months | -0.416 | -1.326, 0.243 | -0.134 | -0.690, 0.422 |
| 12 months | -0.425 | -1.323, 0.226 | -0.141 | -0.698, 0.415 |
| 18 months | -0.444 | -1.331, 0.200 | -0.156 | -0.719, 0.407 |
| 2 years | -0.462 | -1.354, 0.181 | -0.171 | -0.749, 0.407 |
| 3 years | -0.499 | -1.447, 0.169 | -0.200 | -0.830, 0.430 |
| 5 years | -0.573 | -1.779, 0.219 | -0.258 | -1.056, 0.540 |
| 7 years | -0.646 | -2.218, 0.328 | -0.317 | -1.331, 0.698 |
| 8 years | -0.683 | -2.458, 0.393 | -0.346 | -1.478, 0.787 |

All models are weighted for sociodemographic-lifestyle-reproductive factors (i.e., maternal age, parity, marital status, obstetric comorbidity index adapted from Bateman et al., maternal education, maternal gross yearly income, smoking and alcohol use in pregnancy, BMI at conception, history of abortions/miscarriages, folic acid intake, paternal age, paternal education, paternal social benefits, and paternal BMI), maternal psychiatric correlates (i.e., Lifetime History of Major Depression measured in Q1, SCL-5 measured in Q1 and Q3), comedication in pregnancy (i.e., opioid analgesics – ATC code N02A, benzodiazepines/z-hypnotics - ATC codes N05B and N05C, antipsychotics and mood stabilizers - ATC code N05A, and antiepileptics - ATC code N03A)

**Table S18**. Weighted differences in children’s BMI (in kg/m2) up to 8 years of age between prenatal antidepressant exposure groups (n=486 mother-child pairs in those in 4^th^ quartile of polygenic risk score for maternal antidepressant response), overall and by child sex

|  | Antidepressant exposure | | | |
| --- | --- | --- | --- | --- |
|  | Continuers vs discontinuers | | Continuers vs unexposed | |
|  | beta | 95%CI | beta | 95%CI |
| **All children** |  |  |  |  |
| Average | 0.327 | -0.183, 0.838 | -0.133 | -0.471, 0.205 |
| By age |  |  |  |  |
| 1 month | 0.325 | -0.181, 0.831 | -0.134 | -0.469, 0.201 |
| 3 months | 0.321 | -0.177, 0.820 | -0.135 | -0.465, 0.195 |
| 6 months | 0.315 | -0.173, 0.803 | -0.136 | -0.461, 0.188 |
| 8 months | 0.310 | -0.169, 0.788 | -0.138 | -0.458, 0.181 |
| 12 months | 0.304 | -0.167, 0.774 | -0.140 | -0.456, 0.177 |
| 18 months | 0.292 | -0.167, 0.751 | -0.143 | -0.459, 0.173 |
| 2 years | 0.280 | -0.174, 0.734 | -0.146 | -0.469, 0.177 |
| 3 years | 0.257 | -0.206, 0.720 | -0.153 | -0.509, 0.204 |
| 5 years | 0.210 | -0.340, 0.761 | -0.165 | -0.642, 0.311 |
| 7 years | 0.164 | -0.533, 0.861 | -0.178 | -0.811, 0.454 |
| 8 years | 0.141 | -0.642, 0.923 | -0.185 | -0.902, 0.532 |
| **Male** |  |  |  |  |
| Average | 0.158 | -0.640, 0.956 | -0.168 | -0.566, 0.231 |
| By age |  |  |  |  |
| 1 month | 0.161 | -0.627, 0.949 | -0.168 | -0.562, 0.227 |
| 3 months | 0.166 | -0.602, 0.935 | -0.167 | -0.554, 0.220 |
| 6 months | 0.175 | -0.566, 0.916 | -0.166 | -0.544, 0.212 |
| 8 months | 0.184 | -0.531, 0.899 | -0.165 | -0.538, 0.207 |
| 12 months | 0.193 | -0.499, 0.885 | -0.164 | -0.535, 0.206 |
| 18 months | 0.210 | -0.443, 0.863 | -0.163 | -0.538, 0.213 |
| 2 years | 0.228 | -0.398, 0.853 | -0.161 | -0.554, 0.232 |
| 3 years | 0.263 | -0.348, 0.874 | -0.157 | -0.617, 0.302 |
| 5 years | 0.333 | -0.408, 1.074 | -0.150 | -0.815, 0.515 |
| 7 years | 0.404 | -0.599, 1.406 | -0.143 | -1.054, 0.768 |
| 8 years | 0.439 | -0.718, 1.595 | -0.139 | -1.180, 0.901 |
| **Female** |  |  |  |  |
| Average | 0.155 | -0.544, 0.853 | -0.267 | -0.812, 0.277 |
| By age |  |  |  |  |
| 1 month | 0.153 | -0.542, 0.849 | -0.267 | -0.809, 0.275 |
| 3 months | 0.150 | -0.540, 0.840 | -0.266 | -0.804, 0.272 |
| 6 months | 0.145 | -0.539, 0.829 | -0.264 | -0.797, 0.268 |
| 8 months | 0.140 | -0.538, 0.819 | -0.263 | -0.792, 0.266 |
| 12 months | 0.135 | -0.539, 0.810 | -0.261 | -0.788, 0.265 |
| 18 months | 0.126 | -0.544, 0.796 | -0.259 | -0.785, 0.268 |
| 2 years | 0.116 | -0.555, 0.787 | -0.256 | -0.788, 0.277 |
| 3 years | 0.096 | -0.593, 0.786 | -0.250 | -0.812, 0.312 |
| 5 years | 0.057 | -0.726, 0.840 | -0.238 | -0.916, 0.439 |
| 7 years | 0.018 | -0.913, 0.950 | -0.227 | -1.067, 0.613 |
| 8 years | -0.001 | -1.020, 1.018 | -0.221 | -1.153, 0.711 |

All models are weighted for sociodemographic-lifestyle-reproductive factors (i.e., maternal age, parity, marital status, obstetric comorbidity index adapted from Bateman et al., maternal education, maternal gross yearly income, smoking and alcohol use in pregnancy, BMI at conception, history of abortions/miscarriages, folic acid intake, paternal age, paternal education, paternal social benefits, and paternal BMI), maternal psychiatric correlates (i.e., Lifetime History of Major Depression measured in Q1, SCL-5 measured in Q1 and Q3), comedication in pregnancy (i.e., opioid analgesics – ATC code N02A, benzodiazepines/z-hypnotics - ATC codes N05B and N05C, antipsychotics and mood stabilizers - ATC code N05A, and antiepileptics - ATC code N03A)
